# CD3-engaging bispecific antibodies convert human regulatory T cells into cytotoxic effectors

**DOI:** 10.64898/2026.09.23.26363285

**Authors:** Julius J. Michel, Jan H. Frenking, Anna D. Metzler, Antonia Schach, Erin W. Meermeier, Vera Thiel, Niklas Kehl, Tim R. Wagner, Julian Zoller, Sven Cuntz, Cornelius Funk, René Onken, Alanna Kirschner, Stefanie Huhn, Alexander Brobeil, Andreas Trumpp, Mohammad Rahbari, Mathias Heikenwälder, Carsten Müller-Tidow, P. Leif Bergsagel, Hartmut Goldschmidt, Hermann Einsele, Leo Rasche, Marta Chesi, Marc S. Raab, Niels Weinhold, Mirco J. Friedrich

## Abstract

Bispecific T cell engagers (BTCEs) have transformed the treatment of B cell malignancies, but how they remodel non-conventional T cell circuits in patients, and how they should be ideally combined with existing treatment backbones, remains unclear.

Here we show that CD3-engaging BTCEs convert human FOXP3⁺ regulatory T cells (Tregs) into an AP-1-imprinted hybrid cytotoxic state that contributes to tumor clearance. Leveraging longitudinal bone marrow and peripheral blood sampling from patients with newly diagnosed multiple myeloma (NDMM) treated with BTCE-based induction in the phase 2 (GMMG-HD10/DSMM-XX/MajesTEC-5) trial, which achieves uniformly deep, minimal residual disease (MRD)-negative remissions, we integrate single-cell RNA, ATAC and TCR sequencing with functional and metabolic profiling to define the underlying mechanism. BTCE-containing induction drives expansion of cytotoxic CD8⁺ clones and the emergence of a Treg population characterized by GZMA, GNLY and NKG7 expression, enriched cytotoxic gene signatures, and licensed by BATF-AP-1-centered chromatin remodeling. *In vitro*, BTCE-exposed Tregs acquire *bona fide* lytic function, killing tumor cells in a target- and dose-dependent manner and undergoing metabolic rewiring towards an effector-like state while retaining canonical Treg lineage markers. Immunomodulatory drugs (IMiDs) do not initiate but quantitatively amplify and qualitatively reshape this BTCE-driven cytotoxic program, providing a mechanistic rationale for BTCE-IMiD combination regimens. Cytotoxic Treg reprogramming is conserved across BTCEs targeting BCMA, GPRC5D, and CD19 and across multiple myeloma, as well as acute lymphoblastic leukemia.

These findings reveal unexpected plasticity of human Tregs under synthetic T cell engagement and establish cytotoxic Tregs as active effectors, candidate biomarkers and tractable levers for optimizing T cell-redirecting immunotherapy.

**Highlights:**

- Frontline teclistamab (BCMA×CD3) induces deep MRD-negative remissions in newly diagnosed multiple myeloma patients and reshapes the T cell compartment in the phase 2 MajesTEC-5 trial.
- BTCEs convert FOXP3⁺ Tregs into AP-1-imprinted cytotoxic effector cells *in vivo*.
- IMiDs amplify BTCE-driven cytotoxic Treg reprogramming without loss of identity.
- Cytotoxic reprogramming of Tregs is observed across BTCEs and blood cancers.

## Introduction

T cell-redirecting immunotherapies are reshaping the treatment landscape of cancer ^1–4^ and are beginning to expand into autoimmune and other non-malignant diseases ^5–8^. Among these, bispecific T cell engagers (BTCEs) are off-the-shelf antibodies that simultaneously bind a tumor-associated antigen and CD3 on T cells, enforcing synthetic immune synapses and triggering T cell-mediated cytotoxicity independently of classical antigen presentation ^8–10^. BTCEs targeting BCMA or GPRC5D have induced deep and often rapid responses in heavily pretreated patients with relapsed or refractory multiple myeloma ^11–13^, as well as in B cell lymphomas ^14–16^ and acute lymphoblastic leukemia ^17,18^, leading to recent regulatory approvals and rapid clinical adoption. Their favorable activity and feasibility are now propelling BTCEs into earlier disease stages and combination regimens ^19^, including induction therapy in newly diagnosed, transplant-eligible multiple myeloma ^20^. However, responses are not uniformly durable and mechanisms of resistance are emerging; the cellular programs that govern both exceptional efficacy and eventual failure of BTCE therapy remain incompletely understood.

Mechanistic work on BTCE function to date has largely focused on conventional CD8⁺ T cells. We have previously shown that pre-existing T cell states and the capacity to expand CX3CR1⁺ cytotoxic CD8⁺ clones under BTCE pressure critically shape clinical responses, whereas exhausted-like T cell phenotypes are associated with treatment failure ^21^. By contrast, the fate and function of non-canonical T cell subsets under the intense and sustained stimulation imposed by BTCEs are poorly defined. Regulatory T cells (Tregs), characterized by FOXP3 expression and a transcriptional program enforcing immune suppression, are indispensable for peripheral tolerance ^22–28^, yet are frequently co-opted by tumors to restrain anti-tumor immunity ^22–25^. Treg accumulation in the tumor microenvironment correlates with poor prognosis in multiple myeloma and other cancers ^29–34^, and experimental Treg depletion enhances anti-tumor responses in preclinical models ^35–37^. At the same time, studies in autoimmunity, infection and checkpoint blockade have challenged a rigid dichotomy between regulatory and effector T cell fates, and revealed signals of context-dependent Treg plasticity ^38–40^. Whether synthetic T cell engagement by BTCEs stabilizes immunosuppressive Treg circuits, promotes immune escape, or instead repurposes Tregs as active contributors to tumor control is unknown.

Here we combine longitudinal single-cell multi-omics with functional modelling to define how BTCE therapy remodels the diseased T cell compartment. By analyzing bone marrow and peripheral blood from patients with newly diagnosed multiple myeloma treated with BCMA×CD3 BTCE-based induction therapy in the phase 2 MajesTEC-5 trial ^20^, alongside patients receiving standard induction therapy and an independent cohort treated with BCMA×CD3 BTCE monotherapy at relapse ^21^, we map clonal, transcriptional and epigenetic trajectories of major T cell subsets. We show that BTCE therapy not only elicits robust expansion of cytotoxic CD8⁺ clones but also reprograms FOXP3⁺ Tregs into a distinct cytotoxic state marked by induction of GZMA/B, GNLY, NKG7 and TBX21, effector-like metabolic rewiring and BATF-AP-1-centered chromatin remodeling. BTCE-exposed Tregs acquire direct, target-dependent lytic activity against tumor cells while retaining core Treg lineage markers, and this cytotoxic conversion is amplified by immunomodulatory drugs. Cytotoxic Treg reprogramming is conserved across BTCEs targeting BCMA, GPRC5D, and CD19, and across multiple hematologic malignancies. Our data challenge the conventional view of Tregs as passive suppressors in cancer immunotherapy and instead uncover their capacity to be repurposed as active effectors under conditions of synthetic T cell engagement.

## Results

### Frontline BTCE-augmented induction overcomes high tumor burden and reshapes systemic T cell immunity in newly diagnosed myeloma

We have previously shown that, in the relapsed/refractory setting, clonal expansion of effector CD8⁺ T cells is a key immunological correlate of response to BCMA×CD3 bispecific T cell engagers (BTCEs), whereas the emergence of an exhausted-like T cell state is associated with treatment failure ^21^. However, how BTCEs act when deployed as part of first-line induction in treatment-naïve multiple myeloma (MM), and how they interact with current standard backbone regimens, remains unknown.

To address this question, we leveraged longitudinal bone marrow aspirates, peripheral blood and clinical response data from patients enrolled in Majes-TEC-5 (NCT05695508), an ongoing first-line clinical trial that incorporates the BCMA×CD3 BTCE teclistamab into induction therapy. This trial provides a unique view into the immune and clinical consequences of BTCE-based induction in newly diagnosed disease. We obtained paired samples before cycle 1 (pre) and after cycle 3 (post) of BTCE-enhanced induction from 17 patients (teclistamab + daratumumab + lenalidomide + dexamethasone [BTCE-DRd], n=14; teclistamab + daratumumab + bortezomib + lenalidomide + dexamethasone [BTCE-DVRd], n=3). As a standard-of-care (SOC) comparator, we analyzed six patients treated with DRd alone. In addition, we included our previously published cohort of 18 patients with relapsed/refractory MM (RRMM) treated with BCMA×CD3 BTCE monotherapy on NCT03269136 as an external BTCE reference (**Figure 1A, Supplementary Tables 1-2**).

**Figure 1.**
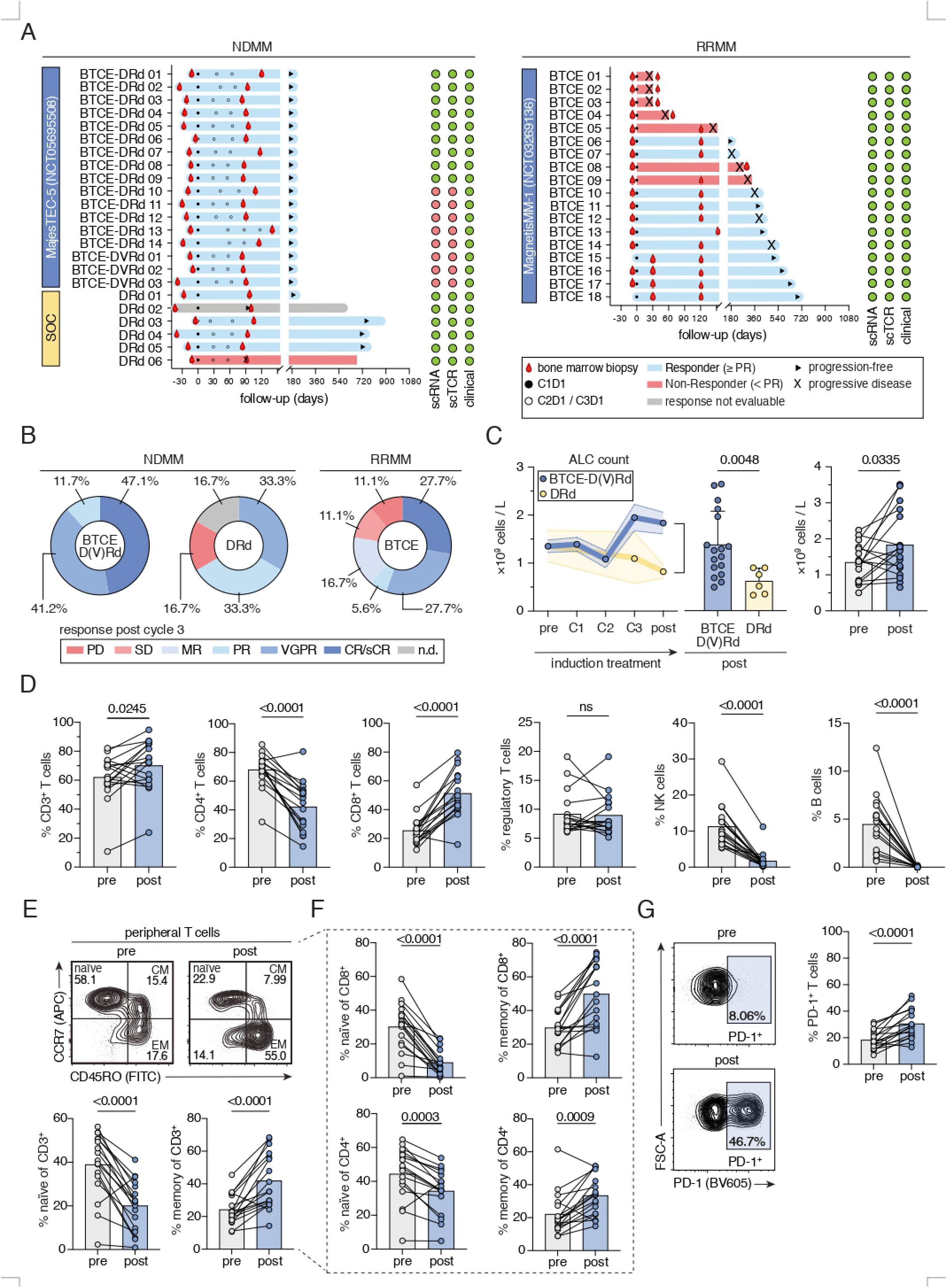
Front-line BTCE therapy intensifies clinical responses and reshapes peripheral T cell composition in newly diagnosed myeloma. **(A)** Swimmer plots depicting treatment timelines, clinical responses, and sample collection points for the newly-diagnosed multiple myeloma (NDMM) (left) and relapsed/refractory multiple myeloma (RRMM) (right) cohorts. BTCE, bispecific T-cell engager. C, cycle. D, day. DRd, daratumumab + lenalidomide + dexamethasone. DVRd, daratumumab + bortezomib + lenalidomide + dexamethasone. PR, partial response. **(B)** Distribution of response categories after cycle 3 for patients with NDMM (BTCE-DRd, n=14; BTCE-DVRd, n=3; DRd, n=6) and patients with RRMM (n=18). Response categories include: PD (Progressive Disease), SD (Stable Disease), MR (Minimal Response), n.d., not done or evaluable. PR (Partial Response), VGPR (Very Good Partial Response), and CR/sCR (Complete Response/Stringent Complete Response). **(C)** Absolute lymphocyte counts (ALC) in peripheral blood over time for BTCE-D(V)Rd and DRd treatment cohorts (left); comparison of post-treatment (after C3) ALC between BTCE-D(V)Rd (n=17) and DRd (n=6) (middle); and paired pre-(before C1) versus post-treatment (after C3) ALC for BTCE-D(V)Rd (n=16) (right). P-values are shown for indicated comparisons and were calculated by Mann Whitney test and Wilcoxon matched-pairs signed rank test, respectively. **(D)** Flow cytometric quantification of major lymphocyte subsets in peripheral blood pre-(before C1) and post-(after C3) BTCE treatment, including the percentage of CD3⁺ T cells, NK cells, and B cells within living PBMCs, the percentage of CD4⁺ T helper cells and CD8⁺ cytotoxic T cells within CD3⁺ T cells, and the percentage of FOXP3⁺ Tregs within CD4⁺ T cells. **(E)** Representative FACS plots and quantification of naïve, central memory (CM), and effector memory (EM) T cells identified by CCR7 and CD45RO expression. **(F)** Quantification of naïve, CM, and EM subsets within CD4⁺ and CD8⁺ T-cell compartments. **(G)** Representative FACS plot and quantification of PD-1 expression on peripheral blood CD3⁺ T cells pre-(before C1) versus post-(after C3) treatment. For panels (D–G), statistical significance was assessed using paired t-tests. Paired data was available for n=17 patients from the BTCE-D(V)Rd cohort.

Clinical responses in the BTCE-augmented induction cohort were both universal and remarkably deep. All Tec-treated patients (17/17, 100%) responded after three cycles (**Figure 1B**), and all were flow-MRD negative at 10^-5^ sensitivity by the post timepoint. Responses continued to deepen with ongoing therapy, resulting in a best overall response of stringent complete response (sCR) in all 17 patients during follow-up. By contrast, in the SOC (DRd) cohort the response rate (partial response or better) after three cycles was 66.7% (4/6); one patient was non-measurable according to International myeloma working group (IMWG) criteria but showed no clinical or radiological progression, and one patient had refractory extramedullary disease. Notably, all DRd-treated patients remained MRD-positive after cycle 3. Strikingly, this disparity in response depth emerged even though the Tec-exposed cohort entered treatment with a markedly greater baseline marrow disease burden (median 60% vs. 25% plasma cell infiltration; p=0.01; **Supplementary Figure S1A-B**), underscoring the capacity of BTCE-containing induction to overcome substantial tumor load.

The incorporation of BTCE into induction therapy was accompanied by systemic expansion of lymphocytes. At baseline (pre), absolute lymphocyte counts (ALC) were comparable between Tec-exposed and DRd patients (median 1.35×10⁹ vs. 1.02×10⁹ cells per liter; p=0.9). By the post timepoint, BTCE-treated patients displayed a significantly higher ALC than DRd-treated controls (median 1.67×10⁹ vs. 0.84×10⁹ cells per liter; p=0.005), and ALC had increased significantly relative to each patient’s own baseline within the Tec cohort (p=0.03; **Figure 1C**). When comparing overall white blood cell and neutrophil counts at the respective time points between both cohorts, no significant differences were found, indicating that BTCE-augmented induction selectively expands lymphocytes rather than globally perturbing hematopoiesis (**Supplementary Figure S1C**).

To define which immune populations contributed to this expansion, we performed multiparametric flow cytometry on paired peripheral blood samples from all 17 Tec-exposed patients (**Supplementary Figure S2A**). Across the induction period, CD3⁺ T cells increased significantly in frequency and absolute number (p=0.0008), an effect largely driven by expansion of CD8⁺ T cells (p<0.0001), while the relative proportions of CD4⁺ T cells, NK cells and B cells declined (**Figure 1D**). Our prior work has implicated effector CD8⁺ T cells as key mediators of clinical response to BTCE therapy in RRMM^21^. In line with a conserved mechanism of action, we observed a significant expansion of effector CD8⁺ T cells in NDMM when BTCE was added to standard induction. Within the CD3⁺, CD4⁺ and CD8⁺ T cell compartments, we observed a marked shift from naïve to effector-memory phenotypes (**Figure 1E-F**). In line with this, Tec-exposed CD3⁺ T cells upregulated surface markers associated with cytotoxic activation (e.g., CD314 and CD29), differentiation (CD45RO), and activation / early exhaustion (PD-1), while concomitantly downregulating lymphoid homing and costimulatory markers characteristic of less differentiated T cells, including CD62L and high-level CD27 expression (CD27^high^) (**Figure 1G and Supplementary Figure S2**), consistent with broad T cell activation and differentiation.

Together, these data show that BTCE-containing induction therapy in NDMM not only drives uniformly deep remissions with early MRD eradication in a high-burden setting, but also reshapes the systemic immune landscape by expanding and activating T cells.

These observations prompted us to ask how such potent clinical and systemic immune effects are encoded at the level of individual T cell states and clonotypes within the bone marrow niche.

### Frontline BTCE therapy remodels T cell clonotypes and installs a cytotoxic effector program in bone marrow T cells

To define how BTCE-based induction therapy reshapes human T cells *in vivo*, we leveraged longitudinal bone marrow aspirates from patients enrolled in Majes-TEC-5 and performed joint single-cell transcriptome and TCR repertoire profiling, enabling us to map, at high resolution, how BTCE-based induction therapy remodels human T cells in patients. We analyzed bone marrow-derived T cells from 9 patients treated with Tec-DRd and six patients receiving SOC DRd, and compared these to 18 patients with RRMM treated with BCMA×CD3 BTCE monotherapy. This design enabled us to distinguish BTCE-driven effects from those of standard induction therapy and to contrast treatment-naïve with heavily pretreated disease (**Supplementary Figure S3**).

By projecting all T cells onto our previously published reference atlas of marrow-derived T cells from MM patients ^41^, we resolved six CD8⁺ T cell subsets, five CD4⁺ subsets and a FOXP3⁺ regulatory T cell (Treg) population shared across cohorts (**Figure 2A, Supplementary Figure S3B-D**). At baseline, NDMM and RRMM patients differed markedly in T cell composition: The bone marrow in NDMM was enriched for CD8⁺ T cells, with a higher proportion of CD8⁺ effector cells, whereas RRMM samples showed reduced overall CD8⁺ T-cell abundance but preserved CD8⁺ effector memory populations, together with a loss of CD4⁺ quiescent cells, potentially related to cumulative effects of age, prior therapies and disease evolution on the T-cell pool. By contrast, within NDMM, baseline T cell composition did not differ significantly between patients subsequently treated with Tec-DRD versus DRD alone, aside from modest inter-patient heterogeneity (**Figure 2B, Supplementary Figure S4A**), indicating that downstream differences reflect treatment rather than pre-existing imbalance.

**Figure 2.**
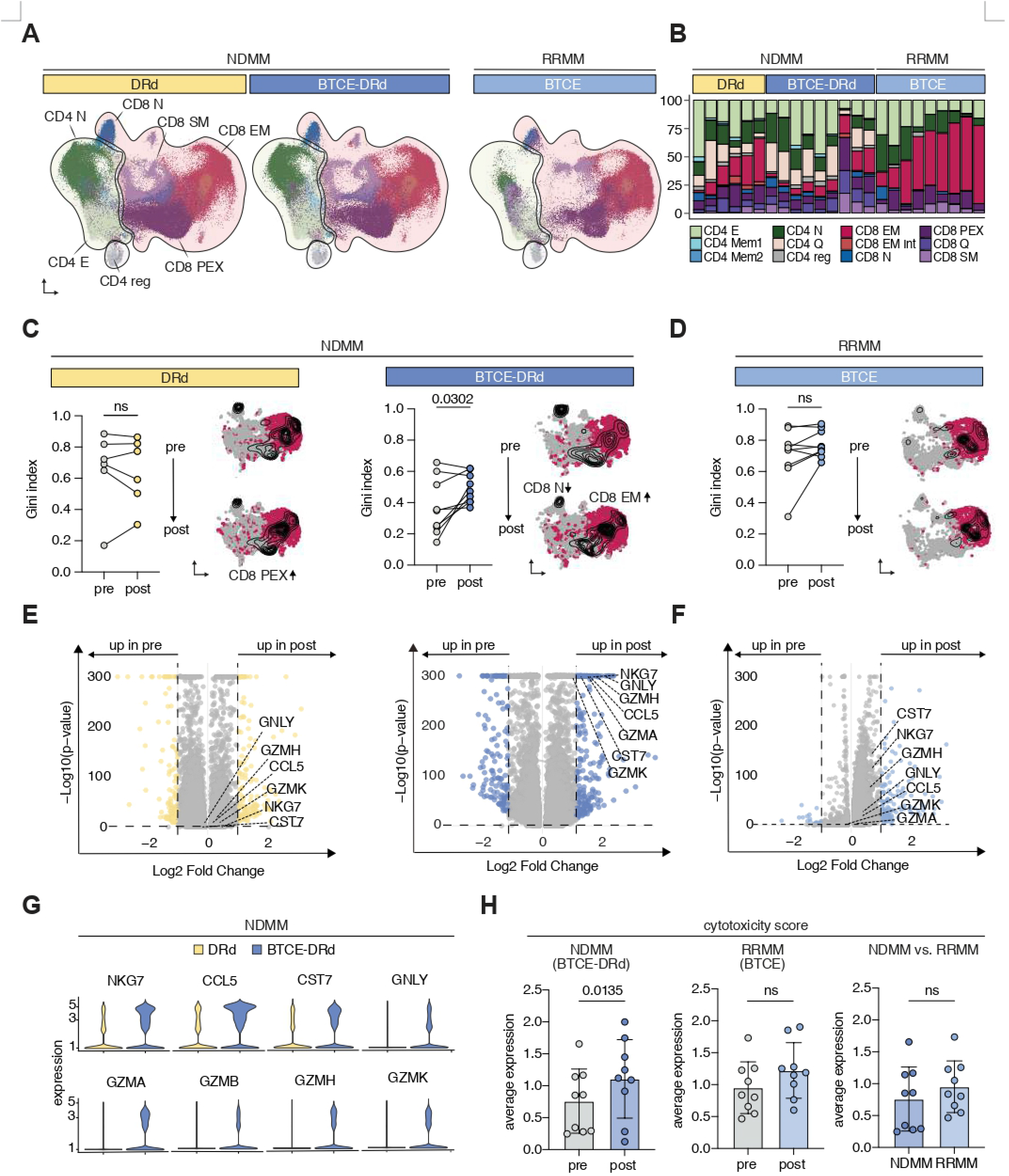
BTCE promotes cytotoxic transcriptional programs and reshapes clonal T cell architecture. **(A)** UMAP visualization of T cell subsets from NDMM and RRMM samples (total 504,969 T cells), annotated into 3 major groups and 12 transcriptionally defined clusters. NDMM, newly-diagnosed multiple myeloma. RRMM, relapsed/refractory multiple myeloma. **(B)** Baseline (pre-therapy / before C1) T-cell cluster distribution across treatment groups, with each bar representing an individual patient. **(C–D)** Gini indices quantifying the spatial distribution of CD8 effector memory (EM) T cells in UMAP space, comparing pre-(before C1) and post-(after C3) treatment time points in the DRd, BTCE-DRd (NDMM), and BTCE (RRMM) cohorts. Each sample was down-sampled to the minimum paired T-cell number. P-values reflect paired t-tests. BTCE, bispecific T-cell engager. DRd, daratumumab/lenalidomide/dexamethasone. **(E–F)** Volcano plots displaying differential gene expression between pre-treatment-exclusive (pre / before C1) and post-treatment-exclusive (post / after C3) T-cell clones across patients. Clones present only pre-therapy were classified as pre; those present only post-therapy as post. P-values were corrected using the Benjamini–Hochberg method. **(G)** Violin plots showing expression of cytotoxicity-associated genes (NKG7, CCL5, CST7, GNLY, GZMA, GZMB, GZMH, GZMK) in NDMM patients treated with DRd (yellow) or BTCE-DRd (blue). **(H)** Cytotoxicity score (see Methods) comparing pre-(before C1) and post-(after C3) therapy samples across NDMM (BTCE-DRd), n=9; RRMM (BTCE), n=9; and baseline NDMM vs. RRMM cohorts. P-values were calculated using paired t-tests.

We next asked whether these phenotypic shifts reflected clonal remodeling. Using the Gini index as a measure of inequality in clonotype size distribution, we found that NDMM patients receiving Tec-DRd exhibited a significant increase in clonal skewing of bone marrow T cells between pre- and post-therapy, indicative of BTCE-induced clonal expansion, whereas DRd-treated NDMM patients did not. A less pronounced increase in clonality was observed in RRMM patients upon BTCE monotherapy (**Figure 2C-D and Supplementary Figure S4B-C**), pointing to a shared clonal response to BTCE across disease stages. To visualize the fate of individual clones, we integrated phenotypic states, TCR clonotypes and longitudinal dynamics into a high-dimensional map across all NDMM and RRMM patients. In NDMM marrows, expanded clones after BTCE were predominantly located within CD8⁺ effector and CD8⁺ quiescent compartments, accompanied by a decrease in clone size within naïve CD8⁺ T cells, consistent with a differentiation trajectory from naïve to effector-like states upon BTCE engagement (**Figure 2C**). RRMM patients showed a similar architecture: expansion of large CD8⁺ effector clones together with contraction of CD8⁺ naïve and CD4⁺ effector clones upon BTCE exposure (**Figure 2D**). DRd-treated patients, however, demonstrated a more pronounced clonal shift towards progenitor-exhausted CD8^+^ phenotypes after three treatment cycles (**Figure 2C**).

To link clonal remodeling to functional states, we performed differential gene expression analyses separately in NDMM and RRMM cohorts, tracking individual T cell clones over time by their clonotype sequences. For each patient, we categorized clones as pre-treatment-exclusive (pre), or post-treatment-exclusive (post). Comparing clones detected only after therapy to those present only before therapy revealed robust induction of canonical cytotoxic effector genes, including *GZMA*, *GZMH*, *GNLY* and *CCL5*, specifically in NDMM patients treated with Tec-DRd (**Figure 2E**). This cytotoxic program was not induced in NDMM patients receiving DRd alone nor in RRMM patients treated with BTCE (**Figure 2E-F**). Consistently, a curated cytotoxic gene signature ^41^ showed strong upregulation between pre and post in NDMM BTCE marrows, but not in DRd-treated NDMM or in RRMM BTCE cohorts (**Figure 2G-H**), identifying the emergence of highly cytotoxic clones as a distinctive feature of frontline BTCE therapy in treatment-naïve disease.

Together, this TCR-resolved analysis of serial bone marrow samples from the ongoing first-line MajesTEC-5 trial in combination with earlier data^21^ demonstrates that BTCE therapy elicits a conserved cytotoxic T cell response across disease stages which was found to be associated with high clinical efficacy and absent in the standard-of-care controls.

### BTCE therapy converts FOXP3⁺ regulatory T cells into hybrid cytotoxic effector Tregs *in vivo*

Having established that frontline BTCE therapy drives the emergence of highly cytotoxic T cell clones, we next asked which compartments encode this heightened effector program. Differential expression analyses stratified by CD8⁺ T cells, CD4⁺ T cells and FOXP3⁺ Tregs revealed that the most pronounced induction of cytotoxic genes between BTCE-treated NDMM and BTCE-treated RRMM unexpectedly occurred within the Treg compartment (**Figure 3C, Supplementary Figure S4D**). To explore this further, we subsetted our integrated dataset of 432,947 marrow-derived NDMM T cells from 15 patients to *FOXP3⁺* Tregs and performed *de novo* clustering. This analysis uncovered an additional Treg cluster that emerged specifically after BTCE-DRd, but not after DRd therapy alone (**Figure 3A**). Cells in this BTCE-associated cluster selectively expressed the cytotoxic effector genes *GZMA*, *GNLY* and *NKG7* while retaining *FOXP3* and *IL2RA* expression and low *IKZF2*, distinguishing them from all other Treg clusters (**Figure 3B-C**). These findings indicate that BTCEs do not merely spare Tregs but actively reprogram a subset of them into a distinct cytotoxic-like state, prompting us to dissect the phenotypic, functional and metabolic underpinnings of this Treg subset in more detail.

**Figure 3.**
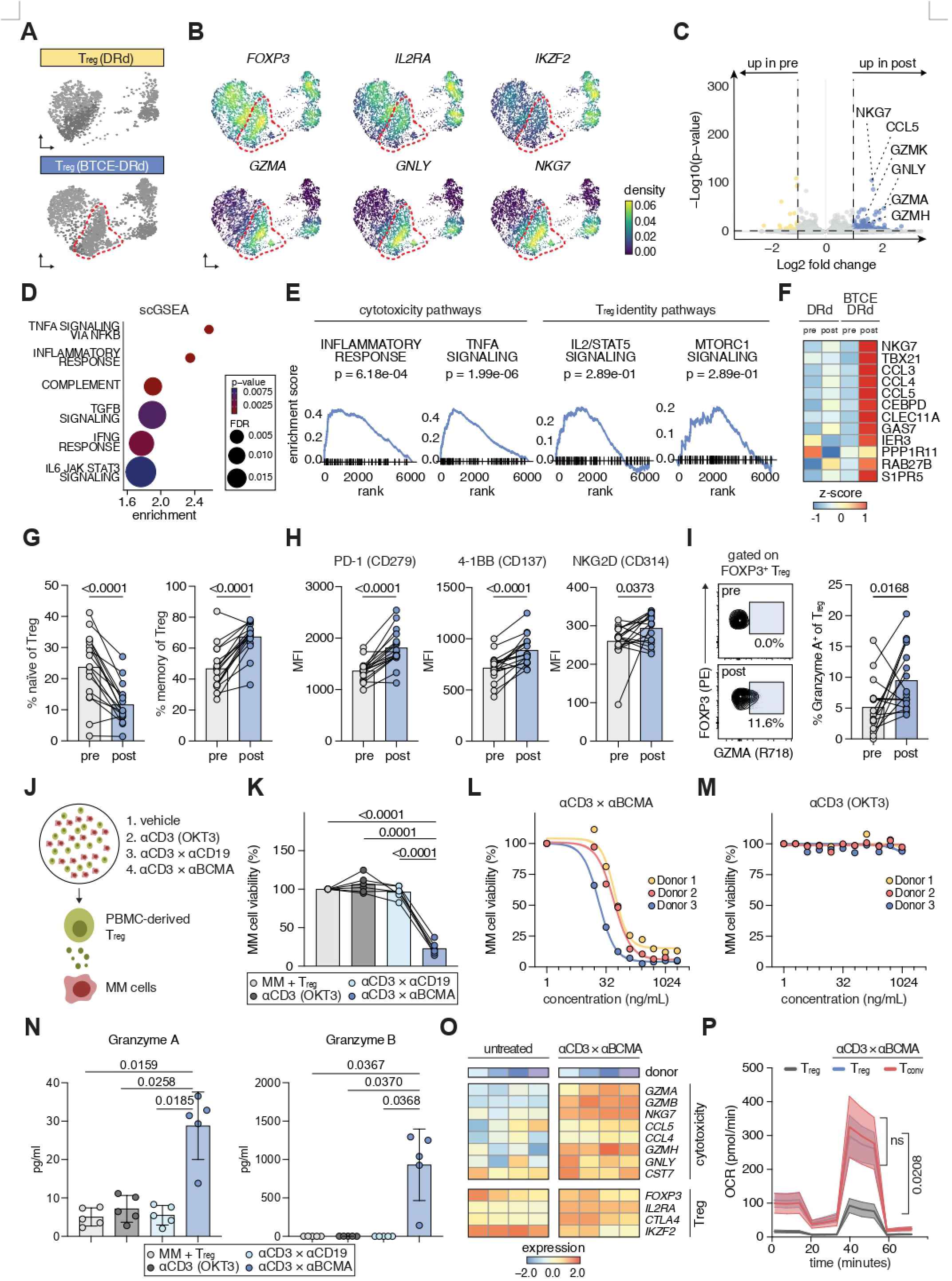
BTCE drives phenotypic, functional, and metabolic reprogramming of Tregs into cytotoxic effector cells. **(A)** UMAP plots showing FOXP3⁺ regulatory T cells (Tregs) in NDMM patients treated with DRd (top) versus BTCE-DRd (bottom). Cytotoxic Tregs are delineated. BTCE, bispecific T-cell engager. DRd, daratumumab + lenalidomide + dexamethasone. NDMM, newly-diagnosed multiple myeloma. **(B)** UMAP expression maps of Treg-identity genes (FOXP3, IL2RA, IKZF2) and cytotoxicity-associated genes (GZMA, GNLY, NKG7) within the Treg compartment. **(C)** Volcano plot showing differential gene expression in Tregs from BTCE-DRd-treated patients comparing post-(after C3) therapy versus pre-(before C1) therapy samples. Upregulated cytotoxicity-associated genes (NKG7, CCL5, GZMK, GNLY, GZMA, GZMH) are highlighted. **(D)** scGSEA dot plot displaying p-values, FDR-adjusted p-values, and enrichment scores for immune pathways upregulated in post-therapy (after C3) Tregs from BTCE-DRd–treated patients. **(E)** Representative enrichment curves for selected pathways, including Inflammatory Response, TNFα signaling via NFκB, IL2/STAT5 signaling, and mTORC1 signaling. **(F)** Heatmap of z-score-normalized expression for literature-defined exTreg signature genes (TBX21, CCL5, NKG7, GAS7, CCL3, CCL4, S1PR5, CEBPD) in DRd and BTCE-DRd Tregs pre-(before C1) and post-(after C3) treatment. The data shown in panels (A)-(F) was obtained by single-cell RNA sequencing of bone marrow samples. **(G)** Flow cytometric quantification of naïve and memory Treg subsets pre-(before C1) versus post-(after C3) treatment. **(H)** Median fluorescence intensity (MFI) of PD-1 (CD279), 4-1BB (CD137), and NKG7 (CD314) on peripheral blood Tregs pre-(before C1) versus post-(after C3) treatment. **(I)** Representative intracellular FACS plots and quantification of FOXP3⁺ Tregs co-expressing Granzyme A (GZMA) in peripheral blood from patients before C1 and after C3 of therapy. Panels **(G–I)** were analyzed using paired t-tests. Data was obtained by flow cytometric analysis of peripheral blood samples. **(J)** Schematic overview of the coculture system. BCMA, B-cell maturation antigen. MM, multiple myeloma. PBMC, peripheral blood mononuclear cells. **(K)** Viability of MM1.S cells after 48-h coculture with healthy donor Tregs treated with Blinatumomab (αCD3×αCD19), OKT3 (αCD3), or Teclistamab (αCD3×αBCMA). Two-way ANOVA (n=6). **(L)** Viability of MM1.S cells after 48-h coculture with Tregs treated with increasing concentrations of Teclistamab (n=3). **(M)** Viability of MM1.S cells after 48-h coculture with Tregs treated with equimolar concentrations of OKT3 (n=3). **(N)** Cytokine secretion (Granzyme A, Granzyme B) after 72-h coculture under the same treatment conditions. Two-way ANOVA (n=6). **(O)** Heatmaps comparing the average expression of cytotoxic and lineage-defining genes between Tregs treated *in vitro* with teclistamab and untreated controls. Data was obtained by bulk RNA sequencing. **(P)** Oxygen consumption rate (OCR) of untreated and Teclistamab-treated Tregs and pan T cells after 48-h coculture, measured using Seahorse XF. Repeated-measures one-way ANOVA (n=5).

We first performed gene set enrichment analysis using HALLMARK pathways from the Molecular Signatures Database. BTCE-exposed Tregs in NDMM showed significant upregulation of pathways associated with cytotoxic and inflammatory signaling, with the highest normalized enrichment scores observed for NF-κB signaling and inflammatory response gene sets. By contrast, hallmark Treg programs such as mTOR signaling and IL-2/STAT5 signaling remained unchanged (**Figure 3D-E**), suggesting that BTCE therapy overlays a cytotoxic transcriptional module onto a largely preserved regulatory core program rather than globally dismantling Treg identity.

Cytotoxic or destabilized Tregs (“exTregs”) have been described in the context of immune checkpoint blockade and chronic inflammation, where they lose suppressive capacity and gain effector functions ^39,42^. To assess whether BTCE-reprogrammed Tregs resemble this exTreg phenotype, we focused on established exTreg-associated markers ^42^, including *TBX21*, *TCF7* and *CCL5*. BTCE-treated NDMM Tregs displayed marked enrichment of exTreg-marker expression between pre and post compared with DRd-treated NDMM controls (**Figure 3F**), indicating that frontline BTCE therapy drives conventional Tregs toward an exTreg-like, cytotoxic state *in vivo*. To validate the Treg reprogramming inferred from our single-cell analyses, we first examined peripheral blood Tregs from BTCE-treated patients by multiparametric flow cytometry (n=17, **Supplementary Figure S5A**). Across all individuals, BTCE exposure drove a consistent shift from a naïve (CD45RA⁺) to a memory (CD45RO⁺) phenotype (**Figure 3G**), indicating that circulating Tregs are pushed into a more differentiated state. In parallel, Tregs upregulated activation and co-stimulatory receptors, including CD137 (4-1BB), CD314 (NKG2D) and PD-1, with particularly pronounced induction of CD137 and NKG2D, consistent with acquisition of effector-like and potentially cytotoxic features (**Figure 3H**). Intracellular cytokine staining of matched pre- and post-therapy samples verified significant induction of Granzyme A in peripheral Tregs (**Figure 3I, Supplementary Figure S5B**), mirroring the induction of *GZMA* expression in bone marrow-associated Tregs at single-cell resolution and indicating that BTCE-driven Treg reprogramming is systemic rather than confined to the tumor niche.

To test whether these phenotypic changes translate into bona fide cytotoxic function, we established an *in vitro* co-culture assay comprising myeloma target cells, bulk T cells and Tregs exposed to distinct stimuli: untreated, anti-CD3 (OKT3) which shares the T cell-engaging binding domain sequence of teclistamab, a non-BCMA-targeting bispecific antibody (blinatumomab), or teclistamab (**Figure 3J, Supplementary Figure S6A-F**). Teclistamab-conditioned Tregs displayed robust cytotoxic activity against myeloma targets, though higher BTCE concentrations were required compared to conventional pan T cells used as a positive control (**Figure 3L, Supplementary Figure S6C-D)**. By contrast, Tregs stimulated with OKT3 or blinatumomab did not mediate detectable killing, indicating that generic T cell activation is insufficient and that target-dependent BTCE engagement is required to elicit Treg cytotoxicity (**Figure 3K)**. Tregs exposed to increasing concentrations of teclistamab, but not OKT3, showed a clear dose-response relationship, with higher BTCE concentrations driving progressively stronger myeloma cell killing (**Figure 3L-M**), demonstrating that the magnitude of Treg cytotoxicity scales with the strength of BTCE-mediated target engagement.

To exclude the possibility that rare contaminating effector T cells accounted for the observed killing, we repeated the co-culture experiments using rigorously sorted CD25⁺CD127^low^ Tregs (**Supplementary Figure S5C**). Even under these more stringent conditions, BTCE stimulation endowed highly purified Tregs with potent cytotoxic capacity, confirming that BTCEs directly reprogram bona fide Tregs rather than merely expanding pre-existing effector contaminants (**Supplementary Figure S6B**). Analysis of co-culture supernatants revealed marked upregulation of classical cytotoxic effector proteins, including Granzyme A and Granzyme B upon BTCE stimulation (**Figure 3N, Supplementary Figure S6E-F**), further corroborating the observed GZMA/GZMB-driven transcriptional phenotype in BTCE-treated patients and that BTCE-conditioned Tregs deploy the same effector molecules typically associated with cytotoxic CD8⁺ T cells and NK cells.

Bulk RNA sequencing of Tregs treated *in vitro* with teclistamab versus untreated controls confirmed a distinct transcriptional shift (**Figure 3O**): BTCE-treated Tregs retained expression of canonical lineage-defining genes such as *FOXP3*, *IL2RA* (CD25), *CTLA4* and *IKZF2* (Helios), yet simultaneously acquired a robust cytotoxic gene signature. This included strong induction of granzyme genes (*GZMA*, *GZMB*), perforin-pathway components, NKG2 family molecules (*NKG7*), chemokines (*CCL4*, *CCL5*) and cytotoxic granule proteins (*GNLY*, *CST7*). We hypothesize that this combination of preserved regulatory core and superimposed effector program argues against complete lineage destabilization and instead supports the existence of a hybrid cytotoxic Treg state induced by BTCE engagement.

Finally, we asked whether this functional and transcriptional reprogramming is accompanied by metabolic remodeling of Tregs. Seahorse extracellular flux analyses of Tregs isolated from the co-culture assay showed that teclistamab-treated Tregs adopt oxygen consumption rate (OCR), extracellular acidification rate (ECAR) and mitochondrial stress profiles that closely resemble those of cytotoxic pan T cells across all time points (**Figure 3P, Supplementary Figure S6G-H**). This convergence on a highly bioenergetic, glycolytic–oxidative state contrasts with the quiescent metabolic profile of non-BTCE-exposed Tregs (**Figure 3P**) and indicates that BTCE engagement orchestrates a coordinated phenotypic, functional, transcriptional and metabolic rewiring of Tregs into effector-like cytotoxic cells.

Together, these data suggest a mechanistic framework for how BTCEs simultaneously mobilize multiple T cell lineages during highly effective disease control.

### AP-1-centered chromatin remodeling licenses the BTCE-induced hybrid cytotoxic Treg state

To define the regulatory logic that enables Tregs to acquire cytotoxic effector function under BTCE pressure, we performed single-cell ATAC-seq on freshly isolated Tregs from the bone marrow of NDMM patients before and after BTCE-DRd therapy. Differential accessibility analysis revealed a gain in chromatin accessibility at the BATF locus after BTCE exposure (**Figure 4A**). BATF, a central component of the AP-1 transcription factor network, typically partners with JUN family members to integrate strong TCR and cytokine signals and to drive effector differentiation, cytotoxic programming and tissue adaptation in conventional T cells under inflammatory conditions ^43,44^. The emergence of an open BATF locus in BTCE-treated Tregs therefore suggested that AP-1-mediated control, classically associated with effector and helper T cell programs, is repurposed in this context to enforce a cytotoxic Treg state.

**Figure 4.**
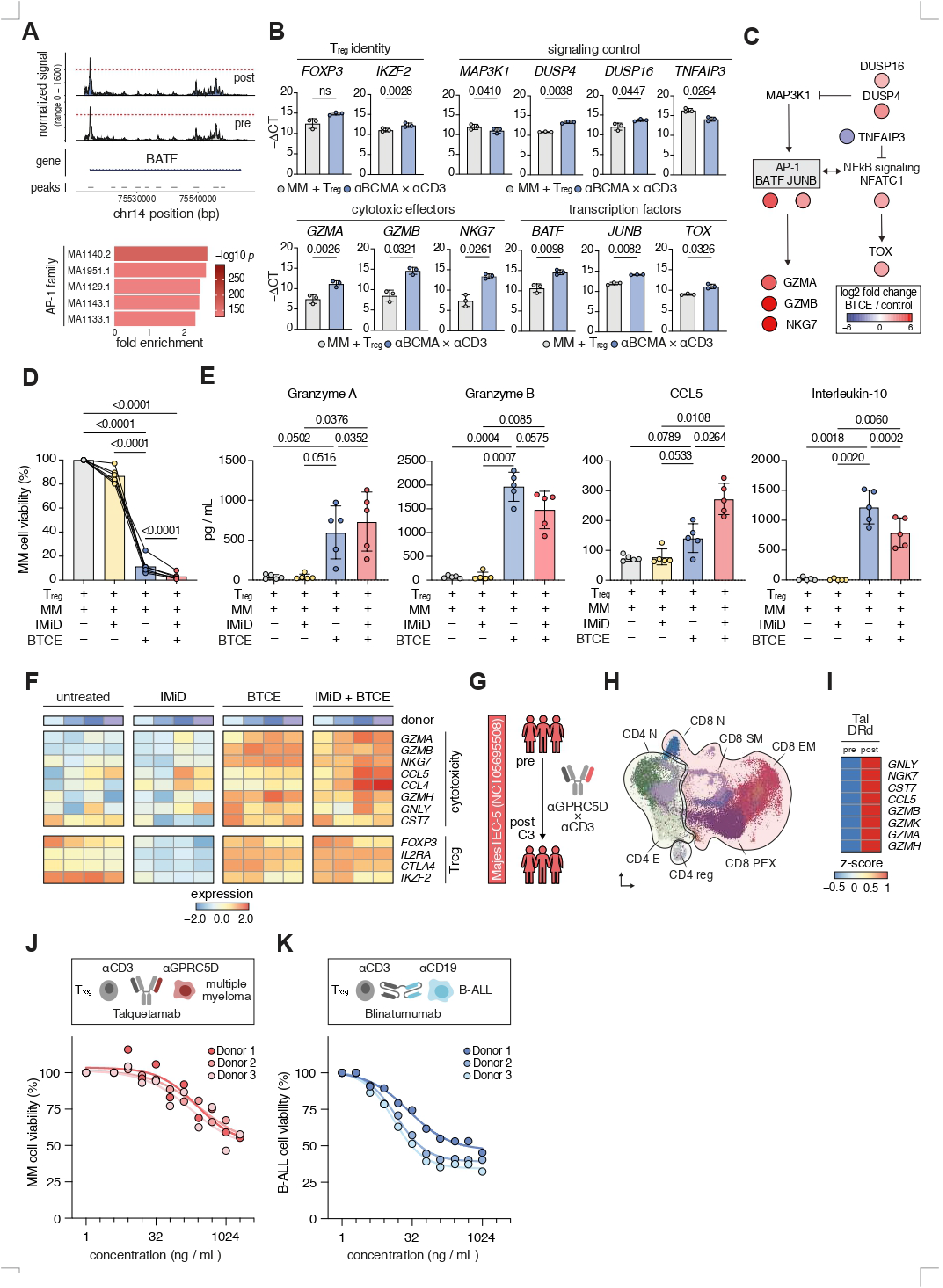
IMiDs potentiate BTCE/AP-1–driven cytotoxic reprogramming of Tregs across bispecific targets and hematologic malignancies. **(A)** Top: ATAC-seq tracks showing differential chromatin accessibility at the BATF locus in Tregs isolated from bone marrow samples obtained post-therapy (after C3) vs. pre-therapy (before C1). Bottom: AP-1 motif enrichment analysis of differentially accessible regions, showing fold enrichment of representative AP-1 family motifs. **(B)** qPCR analysis of Treg gene expression following coculture with MM1.S myeloma cells, with or without Teclistamab (αBCMA×αCD3). Genes are grouped by function: Treg identity (FOXP3, IKZF2), signaling regulators (MAP3K1, DUSP4, DUSP16, TNFAIP3), cytotoxic effectors (GZMA, GZMB, NKG7), and transcription factors (BATF, JUNB, TOX). Paired t-tests were used for comparisons. **(C)** Proposed transcriptional network induced in Tregs by BCMA×CD3 engagement. Node color intensity reflects log₂ fold changes from panel B. (D) Viability of MM1.S cells after 48-h coculture with healthy donor Tregs treated with OKT3, Teclistamab, Lenalidomide, or combinations. Two-way ANOVA (n=5). **(E)** Cytokine secretion (Granzyme A, Granzyme B, CCL5, IL-10) after 72-h coculture under the same treatment conditions. Two-way ANOVA (n=5). **(F)** Heatmaps comparing the average expression of cytotoxic and lineage-defining genes between untreated Tregs and Tregs treated *in vitro* with either the immunomodulatory agent (IMiD) lenalidomide alone, the bispecific T-cell engager (BTCE) teclistamab alone or a combination of both. Data was obtained by bulk RNA sequencing. **(G)** Schematic illustration of n=3 patients with newly-diagnosed multiple myeloma (NDMM) enrolled in the MajesTEC-5 trial (n=2 Arm E; n=1 Arm E1) and undergoing induction therapy with a combination of talquetamab (G protein-coupled receptor class C group 5 member D [GPRC5D]xCD3), daratumumab, lenalidomide and dexamethasone (Tal-DRd). Samples were collected before cycle 1 (C1) (pre) and after cycle 3 (C3) (post). **(H)** UMAP plot showing the distribution of T cell subsets among patients treated with Tal-DRd. Data was obtained by single-cell RNA sequencing of bone marrow samples. **(I)** Heatmap showing the comparison of the z-score-normalized, cytotoxic gene expression of Tregs between the pre-(before C1) and post-(after C3) treatment time points among patients treated with Tal-DRd. Data was obtained by single-cell RNA sequencing of bone marrow samples. **(J)** Viability of MM1.S cells after 72-h coculture with Tregs isolated from healthy donor PBMCs at increasing doses of Talquetamab (αCD3×αGPRC5D) (n=3). **(K)** Viability of Nalm6 cells after 72-h coculture with Tregs Tregs isolated from healthy donor PBMCs exposed to escalating doses of Blinatumomab (αCD3×αCD19) (n=3).

Motif enrichment analysis using JASPAR position frequency matrices (methods) confirmed this hypothesis. Regions that gained accessibility after BTCE therapy were strongly enriched for AP-1 motifs, including BATF-JUNB heterodimer binding sites, indicating coordinated activation of an AP-1-centred regulatory module in reprogrammed Tregs (**Figure 4A**). Additional enrichment of ETS motifs, which are known to cooperate with AP-1 downstream of MAPK signaling, further supported a model in which intense, BTCE-driven CD3 engagement triggers a MAPK-AP-1 axis that rewires Treg chromatin (**Supplementary Figure S6I).** In physiological settings, AP-1/ETS cooperation underlies the transition from quiescent to effector T cell states and stabilizes cytotoxic gene expression in CD8⁺ T cells; our data indicate that a similar circuitry is engaged in Tregs under synthetic TCR stimulation.

To validate these epigenetic findings at the transcriptional level, we analyzed Tregs from *in vitro* co-culture assays by RT-qPCR. In line with increased accessibility at AP-1-associated loci, BTCE-treated Tregs showed strong induction of BATF together with JUNB, its canonical binding partner in the AP-1 complex (**Figure 4B**). We also observed robust upregulation of DUSP4 and DUSP16, dual-specificity phosphatases that attenuate MAPK signaling and are typically induced as feedback regulators in the setting of sustained AP-1 activity. Downstream of this AP-1 activation, BTCE-treated Tregs upregulated hallmark cytotoxic effectors, including *GZMA*, *GZMB* and *NKG7*, as well as increased NF-κB/NFAT/TOX signaling, mirroring the chronic effector program observed *in vivo* and aligning with the known role of BATF-JUN complexes in licensing granule-mediated cytotoxicity (**Figure 4C**).

BTCEs thus co-opt a canonical effector T cell transcriptional circuit and superimpose it onto a FOXP3⁺ lineage, generating a hybrid, AP-1-imprinted cytotoxic Treg state that links strong, antigen-dependent CD3 engagement to the conversion of Tregs from a suppressive into a cytotoxic effector phenotype without loss of core lineage identity.

### IMiDs amplify BTCE/AP-1–driven cytotoxic Treg reprogramming across CD3-engaging bispecific antibodies and hematologic malignancies

Given that BTCE-treated NDMM patients in our cohort received IMiD-containing induction backbones, we next asked whether IMiDs shape the extent and quality of BTCE-driven Treg reprogramming. To address this mechanistically, we extended our *in vitro* co-culture system to include lenalidomide either alone or in combination with BTCE stimulation. Lenalidomide monotherapy did not affect Treg or myeloma cell viability across a broad dose range, indicating no intrinsic cytotoxicity of Tregs in this setting. By contrast, when combined with teclistamab, lenalidomide markedly augmented Treg-mediated tumor killing, resulting in near-complete elimination of myeloma target cells across donors (**Figure 4D**).

ELISA analyses of co-culture supernatants showed that the combination of lenalidomide and BTCE not only increased overall effector output but also qualitatively reshaped the secretome. Co-treatment enhanced secretion of Granzyme A and CCL5, while reducing Granzyme B and Interleukin-10 (IL-10) relative to BTCE alone (**Figure 4E**), suggesting that IMiDs bias BTCE-reprogrammed Tregs toward augmented inflammatory and chemotactic signals and attenuated canonical suppressive cytokines. Bulk RNA sequencing of Tregs from the same cultures revealed that BTCE plus lenalidomide induced a more potent cytotoxic transcriptional program than BTCE alone, with further upregulation of *GZMA*, *GZMB*, *CCL5* and *NKG7* (**Figure 4F**). In both conditions, Tregs retained expression of core lineage genes (*FOXP3*, *IL2RA*, *CTLA4*, *IKZF2*), consistent with the hybrid cytotoxic Treg state defined above. In contrast, lenalidomide monotherapy downregulated Treg identity markers without inducing a cytotoxic module, indicating that IMiDs alone destabilize Treg lineage programs but require concomitant BTCE engagement to convert this destabilization into productive cytotoxic differentiation (**Figure 4F**). We next asked whether cytotoxic Treg reprogramming was specific to BCMA targeting or represented a broader property of CD3-engaging bispecific antibody therapy. Analysis of scRNA-seq data from NDMM patients treated with talquetamab (GPRC5D×CD3) in combination with IMiD-containing induction in another study arm of the MajesTEC-5 trial showed that post-treatment Tregs acquired the same cytotoxic transcriptional signature seen with teclistamab (**Figure 4G-I, Supplementary Figure S7A-B**), indicating that this phenotype is not epitope-restricted within myeloma. In parallel, talquetamab-treated Tregs in co-culture with multiple myeloma cells displayed dose-dependent killing of myeloma targets (**Figure 4J**), corroborating that BTCE-mediated Treg cytotoxicity can be elicited against distinct tumor antigens. To test whether this phenomenon extends beyond myeloma, we also evaluated Treg cytotoxicity in response to blinatumomab (CD19×CD3) against acute lymphoblastic leukemia cells (B-ALL). In this setting, Tregs acquired potent, dose-dependent cytotoxic activity (**Figure 4K**), demonstrating that Treg reprogramming into cytotoxic effectors is a generalizable feature of CD3-engaging bispecific antibodies across hematologic malignancies, with IMiDs serving as quantitative enhancers rather than absolute requisites of this response.

These findings provide a unifying framework for the context specificity observed in our *in vivo* flow cytometry and single-cell analyses. The uniquely strong cytotoxic Treg program detected in BTCE-treated NDMM marrows, but not in RRMM patients receiving BTCE monotherapy, is more parsimoniously explained by the presence or absence of IMiD co-therapy than by disease stage alone. BTCEs appear to provide the essential antigen-dependent CD3 signal that installs an AP-1-imprinted cytotoxic module in FOXP3⁺ Tregs, while IMiDs amplify and shape this program by modulating T cell transcriptional control and cytokine production. In combination, these agents synergistically convert Tregs from a predominantly suppressive population into hybrid cytotoxic-regulatory effectors, a response conserved across different BTCE targets and hematologic entities. Thus, we hypothesize that what initially appeared as a NDMM-restricted phenomenon likely reflects the broader principle that BTCE-IMiD combinations, rather than disease stage per se, are the critical determinants of the magnitude and character of Treg cytotoxic reprogramming.

Collectively, our clinical, single-cell, functional and epigenomic data show that CD3-engaging BTCE therapy, particularly in combination with IMiDs, remodel the anti-tumor immune landscape by co-opting a canonical effector T cell program in FOXP3⁺ Tregs. Our findings establish AP-1-imprinted cytotoxic Treg reprogramming as a generalizable mechanism of action across BTCE platforms and hematological malignancies, providing a mechanistic basis for biology-informed rational design and optimization of future T cell-redirecting regimens.

## Discussion

BTCEs have rapidly emerged as a transformative class of T cell–redirecting agents, yet their impact on non-canonical T cell compartments has remained largely unexplored ^21^. Here we show that, in the setting of BTCE-based induction therapy for newly diagnosed myeloma, human FOXP3⁺ regulatory T cells are not passive bystanders but are actively reprogrammed into cytotoxic effector cells that support tumor clearance. Across longitudinal single-cell transcriptomic, epigenomic and TCR profiling, together with in-depth functional and metabolic assays, we identify a distinct population of cytotoxic Tregs that arises under BTCE pressure, marked by induction of GZMA/B, GNLY, NKG7, TBX21 and an AP-1-centred chromatin program, while retaining core Treg lineage features. This hybrid state is reinforced by immunomodulatory drugs such as lenalidomide and is conserved across BTCEs, target antigens and hematologic malignancies, suggesting cytotoxic reprogramming of Tregs as a generalizable feature of T cell-engaging therapy rather than a context-restricted epiphenomenon.

Our data extend current models of BTCE mechanism of action, which have so far focused almost exclusively on conventional CD8⁺ T cells. In keeping with prior work, teclistamab-based induction in transplant-eligible newly diagnosed multiple myeloma elicits robust clonal expansion of CD8⁺ effector cells and a shift of pre-existing naïve clones toward cytotoxic effector states, accompanied by deep and universal MRD negativity after only three cycles of therapy ^20^. Against this backdrop, we uncover that the most pronounced induction of cytotoxic gene expression under BTCE pressure occurs not in CD8⁺ T cells but within the FOXP3⁺ Treg compartment. Tregs in BTCE-treated patients upregulate a broad cytotoxic program, acquire memory-like and activation phenotypes and perform active target cell lysis that is absent in patients receiving standard-of-care induction or in relapsed/refractory patients treated with BTCE monotherapy. These observations challenge the canonical view that Tregs in myeloma predominantly restrain tumor immunity and instead indicate that, under conditions of intense synthetic T cell engagement in combination with IMiDs, Tregs can be repurposed as active participants in anti-tumor responses.

Mechanistically, BTCE-induced Treg reprogramming is distinguished by the acquisition of effector-like functions without complete erosion of Treg lineage identity. BTCE-exposed Tregs retain *FOXP3*, *IL2RA*, *CTLA4* and *IKZF2* expression, yet simultaneously engage a potent cytotoxic transcriptional module and remodel their metabolism towards an effector-like, highly bioenergetic state that resembles CD8^+^ T cells. Multiome analyses implicate AP-1-driven chromatin remodeling as a central regulatory hub in this process: teclistamab-treated Tregs exhibit increased accessibility at the BATF locus, broad enrichment of AP-1 motifs and coordinated induction of BATF-JUNB and downstream cytotoxic target genes. *In vitro*, these changes endow Tregs with bona fide lytic activity against tumor cells in a strictly target-dependent and dose-dependent manner. Although their killing potency does not reach that of CD8⁺ T cells, they secrete a comparable repertoire of classical granule-derived effector molecules. The requirement for antigen-specific BTCE engagement, together with the preservation of FOXP3, suggests that cytotoxic Tregs occupy a distinct, AP-1-imprinted hybrid state rather than simply representing unstable or “failed” Tregs drifting toward conventional effector fates or engaging in unspecific cytotoxicity.

The clinical backbone in which BTCEs are deployed appears to further shape this Treg plasticity. Immunomodulatory drugs are a cornerstone of myeloma therapy and have pleiotropic, context-dependent effects on T and other immune cells ^45–48^. In our system, lenalidomide alone destabilizes Treg identity markers without inducing a cytotoxic program, whereas its combination with BTCEs amplifies Treg-mediated tumor killing and reshapes the effector secretome by enhancing GZMA and CCL5 while attenuating GNLY, GZMB and IL-10. These findings provide a mechanistic rationale for the exceptional depth of response observed with the BTCE-IMiD-based induction regimen of the MajesTEC-5 trial and suggest that IMiDs act not only by modulating conventional effector T cells and myeloma cells, but also by tuning the magnitude and quality of Treg cytotoxic reprogramming. Most recently, Duell and colleagues showed that higher Treg abundance in the peripheral blood was associated with inferior responses to the CD19-targeting BTCE blinatumomab in relapsed/refractory acute lymphoblastic leukemia, likely reflecting BTCE-induced Treg activation with enhanced IL-10 production and suppression of effector T cell function^49^. Although this appears to contrast with our finding that Tregs can contribute to cytotoxic tumor control, a key distinction is the therapeutic context: unlike blinatumomab monotherapy, the BTCE regimens studied here incorporate immunomodulatory drugs, with lenalidomide profoundly reshaping Treg function and favoring cytotoxic reprogramming over suppressive activity. This highlights that Treg responses to BTCEs are context dependent and can be steered toward anti-tumor effector states by rational combination therapy. More broadly, the observation that cytotoxic Tregs emerge under BTCEs targeting BCMA, GPRC5D, and CD19, and across multiple myeloma, and acute lymphoblastic leukemia, argues that Treg reprogramming is a fundamental and conserved consequence of CD3-engaging bispecific therapy. This generalizability raises the possibility that harnessing or steering Treg plasticity could improve efficacy across diverse T cell-engaging platforms.

The emergence of cytotoxic Tregs has important implications for both efficacy and safety of T cell-redirecting therapies. In principle, Treg reprogramming could simultaneously relieve local immunosuppression and add an additional cytotoxic effector pool, thereby contributing to the rapid debulking and deep remissions we observed in the analyzed trial participants. At the same time, sustained perturbation of Treg identity raises concerns about loss of peripheral tolerance and immune-mediated toxicity, particularly as BTCEs are moved into earlier disease stages, maintenance settings and non-malignant indications. In our study, BTCE-treated Tregs maintain expression of canonical regulatory genes and patients do not exhibit overt autoimmunity within the follow-up available, suggesting that the cytotoxic Treg state may be transient and at least partly self-limiting. However, longer observation and more granular correlation of cytotoxic Treg abundance and phenotype with immune-related adverse events, including infections, cytopenias and organ-specific inflammation, will be required to define whether this plasticity can be uncoupled from toxicity or, conversely, whether dysregulated Treg reprogramming contributes to adverse outcomes.

Our work has several limitations. The patient cohorts studied are of modest size and differ in the dosing schedule of the treatment backbone, constraining our ability to disentangle the relative contributions of BTCE exposure, disease evolution and prior therapy to Treg plasticity. We did not directly manipulate Tregs *in vivo*, for example, by depleting or selectively blocking AP-1 signaling in Tregs during BTCE therapy - so the quantitative contribution of cytotoxic Tregs to tumor control relative to CD8⁺ effector cells remain inferential. In addition to that, our data do not address the durability or reversibility of the cytotoxic Treg state once BTCE pressure is removed.

Finally, our results suggest that depletion (anti-CD38) and reprogramming (BTCE) may be complementary acting on partially overlapping regulatory compartments to favor effector function consistent with the potent clinical activity observed for the teclistamab-daratumumab doublet ^50,51,52^. In our study, BTCE-DRd was associated with the emergence of an additional FOXP3⁺ Treg state that was not observed with DRd alone and that preserved FOXP3/IL2RA while acquiring a cytotoxic effector program (e.g., GZMA, GNLY, NKG7), consistent with a phenotype conversion rather than simple Treg sparing. A potential explanation is that daratumumab preferentially prunes an activated CD38high suppressive Treg pool, lowering suppressive tone, while teclistamab delivers antigen-anchored CD3 engagement that can repurpose remaining FOXP3⁺ cells into an AP-1-imprinted cytotoxic hybrid state; in our functional systems, lenalidomide amplifies this BTCE-driven reprogramming, indicating that the induction backbone can shape both the magnitude and quality of Treg plasticity. Definitive mechanistic dissection in the frontline setting will require prospective factorial designs incorporating Tec-only and Dara-only comparators coupled to CD38-resolved longitudinal Treg profiling to quantify the relative contributions of regulatory cell depletion versus cytotoxic reprogramming, which we were not able to infer from the control cohorts available in the present study. Despite these caveats, our findings reposition Tregs as dynamic participants in T cell-redirecting therapy and open several avenues for therapeutic innovation. Prospective studies should determine whether the frequency, transcriptional profile or chromatin state of cytotoxic Tregs predicts depth or durability of response, resistance, or toxicity, and whether these features can serve as biomarkers for tailoring BTCE dosing and combination partners. Mechanistic experiments in suitable *in vivo* models will be needed to test whether selectively enhancing beneficial aspects of Treg cytotoxic reprogramming, or stabilizing regulatory features while preserving cytotoxic potential, can widen the therapeutic window. Conversely, if excessive or dysregulated Treg plasticity emerges as a driver of toxicity, strategies to temper AP-1 activity or to compartmentalize Treg reprogramming could be explored. More broadly, our data suggest that future designs of T cell-engaging agents and their combinations should explicitly consider the diverse fates not only of CD8⁺ effector cells but also of Tregs and other unconventional T cell subsets. By understanding and harnessing this unexpected plasticity, it may become possible to engineer T cell-redirecting regimens that mobilize the full breadth of the human T cell repertoire while maintaining immune homeostasis.

### Methods Human subjewcts

#### Patients with newly-diagnosed multiple myeloma

This study included a total of 26 treatment-naïve patients with newly-diagnosed multiple myeloma. Patient and disease characteristics are summarized in **Supplementary Tables 1-2**. Twenty patients had received a BTCE-enhanced induction therapy as part of the prospective, multi-cohort phase 2 clinical trial (GMMG-HD10/DSMM-XX/MajesTEC-5) (NCT05695508), including 14 patients treated with Tec-DRD (n=5 Arm A; n=6 Arm A1; n=3 Arm D), three patients treated with Tec-D-VRD (Arm B) and three patients treated with Talq-DRD (n=2 Arm E; n=1 Arm E1). Six patients with NDMM had been primarily treated with standard-of-care DRD at Heidelberg University Hospital. To ensure consistency and comparability of sample quality, processing time, and pre-analytical handling, only patients treated and sampled at Heidelberg University Hospital were included in the present analysis. Longitudinal, paired bone marrow aspirate and peripheral blood samples were collected prior to treatment initiation (pre) and after three cycles of treatment (q4w) (post), unless otherwise stated, and immediately processed as described below. Individual sampling time points, course of therapy, follow-up and available data sets are illustrated in **Figure 1A**. All participants provided written informed consent for sample collection, tissue sequencing, and review of medical records. The study was conducted in accordance with the principles of the Declaration of Helsinki and the Belmont Report. Ethical approval for the collection, functional testing, and sequencing of human samples as well as for clinical data analysis was granted by the Ethics Committee of Heidelberg University Medical Faculty (reference numbers S-096/2017, S-777/2024).

#### Patients with relapsed/refractory multiple myeloma

As additional control, our previously published dataset - including a total of 18 patients with relapsed/refractory multiple myeloma enrolled in a prospective clinical trial (NCT03269136) and undergoing BCMAxCD3 BTCE monotherapy - was reanalyzed ^21^. Samples were collected prior to treatment initiation (pre), after cycle 1 (post/early) and after cycle 4 or at relapse, whichever occurred first (post/late) (**Figure 1A**).

#### Processing of human bone marrow samples

Bone marrow aspirates were 1:1 diluted in preparation buffer (PBS with 0.1% BSA and 2 mM EDTA), and mononuclear cell separation was performed by density centrifugation (Bicoll separating solution, Biochrom) with diluted bone marrow cells (centrifugation 20 min, 1300g). Cells were carefully aspirated and washed with preparation buffer (centrifugation 5 min at 470g). Red blood cells were lysed using RCL buffer (155 mM NH4Cl, 10 mM KHCO3, 0.1 mM EDTA) for 10 min at room temperature and bone marrow cells were washed (centrifugation 5 min, 470g) and resuspended in preparation buffer. Malignant plasma cells were freshly isolated using CD138 MicroBeads, human (Miltenyi Biotec) according to the manufacturer’s instructions and frozen in 90% FBS (Sigma-Aldrich) supplemented with 10 % DMSO and stored in liquid nitrogen until further use. Non-plasma bone marrow mononuclear cells were frozen after cell counting at 1 × 10^7^ cells per aliquot in 90% FBS (Sigma-Aldrich) supplemented with 10 % DMSO and stored in liquid nitrogen until further use.

#### T cell immunophenotyping by spectral flow cytometry

Patient PBMCs were thawed in X-vivo 15 media with 0.04% ul Benzonase. For surface marker phenotyping, cells were washed in PBS, followed by live dead staining with Fixable Viability Stain 440UV (BD, Cat# 566332) for 15min at 4°C. Samples were then washed with PBS and subsequently stained for surface markers (CD4 - BUV395 (BD, clone: SK3, Cat# 563550), CD34 - BUV496 (BD, clone: 581, Cat# 749903), CD33 - BUV563 (BD, clone: WM53, Cat# 741369), CD314 - BUV615 (BD, clone: 1D11, Cat# 751232), CD19 - BUV661 (BD, clone: SJ25C1, Cat# 741604), CD45RA - BUV737 (BD, clone: HI100, Cat# 612846), CD45 - BUV805 (BD, clone: HI30, Cat# 612891), CD29 - BV421 (BD, clone: TS2/16, Cat# 568993), LAG-3 - BV510 (Biolegend, clone: 11C3C65, Cat# 369318), CD8 - BV570 (Biolegend, clone: RPA-T8, Cat# 301038), CD56 - BV605 (Biolegend, clone: 5.1H11, Cat# 362538), PD-1 - BV650 (BD, clone: EH12.1, Cat# 564104), CD94 - BV711 (BD, clone: HP-3D9, Cat# 743952), CD137 - BV750 (BD, clone: 4B4-1, Cat# 569693), CD62L - BV786 (BD, clone: SK11, Cat# 565311), CD45RO - FITC (Biolegend, clone: UCHL1, Cat# 304204), CD69 - RB613 (BD, clone: FN50, Cat# 571124), CD27 - BB700 (BD, clone: M-T271, Cat# 566449), CD138 - RB780 (BD, clone: MI15, Cat# 569110), CD127 - PE (Biolegend, clone: A019D5, Cat# 351304), CD25 - PE-CF594 (BD, clone: M-A251, Cat# 562403), CXCR3 - PE-Cy7 (BD, clone: 1C6, Cat# 560831), CCR7 - APC (BD, clone: 2-L1-A, Cat# 566762), CD3 - APC-Cy7 (BD, clone: SK7, Cat# 557832)).

To measure intracellular cytokine production, PBMCs were restimulated with PMA(50ng/ml) and Ionomycin (500ng/ml) to induce a recall response and incubated with BD GolgiPlug™ Protein Transport Inhibitor (Containing Brefeldin A) (BD, #555029) for 6hr to induce intracellular cytokine accumulation within the golgi vesicles. Following live dead staining with Fixable Viability Dye eFluor™ 780 (eBioscience™, Cat# 65-0865-18) for 15min at 4°C surface markers (CD3 - BV786 (BD, clone: SK7, Cat# 563800), CD4 - BUV395 (BD, clone: SK3, Cat# 563550), CD8 - BV605 (BioLegend, clone: SK1, Cat# 344742), CD25 - BV650 (BD, clone: M-A251, Cat# 563719), CD127 - AF647 (BD, clone: HIL-7R-M21, Cat# 558598)) were stained for 30min at 4°C. Cells were washed and permeabilized with the eBioscience™ Foxp3 / Transcription Factor Staining Buffer Set (Invitrogen™, #00-5523-00) following manufactures protocol. Intracellular cytokines (Granzyme A - R718 (BD, clone: CB9, Cat# 566976), Granzyme B - RB780 (BD, clone: GB11, Cat# 568705), Granulysin - AF488 (BD, clone: RB1, Cat# 558254), CCL5 - BV421 (BD, clone: 2D5, Cat# 564754), IFNy - RB613 (BD, clone: B27, Cat# 571095)) were stained together with anti-FoxP3 - PE (BD, clone: 236A/E7, Cat# 560852) antibody for 30min at room temperature in the dark. Following staining samples were washed and resuspended in FACS buffer and analyzed on the Cytek^®^ Aurora.

#### Fluorescence-activated cell sorting (FACS) of CD45^+^ CD3^+^ T cells

To isolate T cells for single-cell analyses, samples were thawed in a water bath at 37°C, transferred to RPMI cell culture medium with 10% FBS and 0.02% benzonase, counted, divided into eight aliquots per patient sample and centrifuged at 400 g for 5 min at 4°C. The subsequent steps were performed on ice. Cells were resuspended in FACS buffer (1x PBS with 0.4% BSA), washed and stained with a sample-specific TotalSeq^TM^ hashtag antibody (BioLegend), a life/dead marker (eFluor 780) and fluorescence antibodies for flow cytometry (CD45 PerCP-Cy5.5, CD3 FITC and CD38 APC) prior to pooling. After incubation, cells were washed, resuspended in FACS buffer and subjected to FACS.

#### Fluorescence-activated cell sorting (FACS) of CD4^+^ CD127^low^ CD25^+^ regulatory T cells

Following PBMC isolation, cells were stained with anti CD3 - BV421 (BD, clone: SK3, Cat# 563789), anti CD4 - BUV395 (BD, clone: SK3, Cat# 566923), anti CD25 - PE (BD, clone: M-A251, Cat# 555432), anti CD127 - AF647 (BD, clone: HIL-7R-M21, Cat# 558598), and Fixable Viability Dye eFluor™ 780 (eBioscience™, Cat# 65-0865-18), washed in PBS and resuspended in FACS buffer. Cells were sorted on a on a BD Aria Fusion™ system for CD4^+^ CD127^low^ CD25^+^ regulatory T cells and subsequently cocultured as described below.

#### Isolation of CD4^+^ CD127^low^ CD25^+^ regulatory T cells

For functional *in vitro* experiments and combined single-cell gene expression analysis and ATAC sequencing, regulatory T cells were isolated. For *in vitro* experiments, buffy coats were diluted 1:1 in PBS and mononuclear cell separation was performed by density centrifugation (centrifugation 35min, 450g) with Ficoll-Paque™ PLUS density gradient media (Cytiva, #171440039). Cells were carefully aspirated and washed with PBS twice (centrifugation 10min, 300g). Subsequent isolation of Tregs using the EasySep^TM^ human CD4^+^ CD127^low^ CD25^+^ regulatory T cell kit (Stemcell Technologies, Cat# 18063) following manufacturer’s instructions was performed. For combined single-cell gene expression analysis and ATAC sequencing, patient samples were thawed and EasySep^TM^ isolation of regulatory T cells was performed. To assess the proportion of viable regulatory T cells before sample processing, cells were stained with anti CD3 - BV421 (BD, clone: SK3, Cat# 563789), anti CD4 - BUV395 (BD, clone: SK3, Cat# 566923), anti CD25 - PE (BD, clone: M-A251, Cat# 555432), anti CD127 - AF647 (BD, clone: HIL-7R-M21, Cat# 558598), and Fixable Viability Dye eFluor™ 780 (eBioscience™, Cat# 65-0865-18), followed by flow cytometry analysis.

### Single-cell transcriptome / V(D)J / ATAC analyses

#### Single-cell sequencing and data preprocessing

For single-cell RNA (scRNA), and V(D)J analyses, viable CD45⁺CD3⁺ cells were isolated using fluorescence-activated cell sorting on a BD Aria Fusion™ system as described above and immediately processed for subsequent single-cell RNA and TCR sequencing (**Supplementary Figure 1**). Single-cell capture, reverse transcription, and library preparation were performed on the Chromium platform (10x Genomics) using the Single Cell 5′ Reagent Kit v2, following the manufacturer’s protocol, with 40,000 cells loaded per channel. Library quality was assessed, and final libraries were paired-end sequenced (26 and 92 bp) on an Illumina NovaSeq 6000 S2 lane. Raw sequencing data encompassing gene expression, V(D)J, and cell hashing information were processed using Cell Ranger (v7.1) with the ‘multi’ command, referencing the GRCh38 genome (2020-A) and V(D)J reference (v7.1.0).

For combined single-cell gene expression analysis and assay for transposase-accessible chromatin (ATAC) sequencing CD4^+^ CD127^low^ CD25^+^ regulatory T cells were isolated from patient BMNCs followed by low input nuclei isolation (10X Genomics, Nuclei Isolation for Single Cell Multiome ATAC + GEX Sequencing protocol CG000365 Rev C). Recovered nuclei were used for transposition and GEM generation on a Chromium Next Gem Chip J with the Chromium X Series according to the Chromium Next GEM Single Cell Multiome ATAC + Gene Expression User guide (10X Genomics, CG000338 Rev G, Cat# PN-1000285, PN-1000230, PN-1000212, PN-1000215). Subsequent cleanup and library preparation were performed according to the manufacturer’s guidelines. The quality of scATAC libraries was assessed on a TapeStation D1000 (Agilent, Cat# 5067-5586, 5067-5583, 5067-5602, 5067-5582), and single-cell gene expression libraries on a TapeStation D5000 (Agilent, Cat# 5067-5590, 5067-5589, 5067-5588). Sequencing was performed on an Illumina NovaSeq 6000 using a paired-end 50/8/24/49 cycle configuration on an S4 flow cell.

#### Quality control and normalization

Demultiplexing of single-cell data was performed using the HTODemux function in Seurat (v5.0.1), classifying cells as singlets, doublets, or unassigned based on hashtag oligos. Heterotypic doublets were identified with the scDblFinder package and, along with hashtag-identified doublets, removed from further analysis. Cells lacking a productive alpha/beta TCR were excluded. All datasets were normalized using SCTransform and mapped to a reference dataset of pre-annotated bone marrow scRNA-seq profiles from newly diagnosed MM patients. This step facilitated removal of contaminating CD3⁻ cells, likely arising from sorting artifacts. Mapping was performed using the MapQuery function in Seurat, following the SCTransform v2 vignette (https://satijalab.org/seurat/articles/sctransform_v2_vignette.html). Only cells with >200 and <3000 detected genes, <10% mitochondrial content, and identified as singlet CD3⁺ T cells were classified as high-quality and retained for analysis.

#### Differential expression and pathway enrichment

Differential gene expression was performed using Seurat’s FindMarkers() with the MAST hurdle model on normalized data. P-values were adjusted for multiple hypothesis testing using the Benjamini–Hochberg method. Pathway-level changes were assessed using single-cell gene set enrichment analysis (scGSEA) applied to curated Hallmark pathways. Enrichment scores, raw p-values, and FDR-adjusted p-values were calculated for each pathway, and running enrichment curves were generated to visualize differences between pre- and post-therapy Tregs for selected pathways.

#### Module score of T cell cytotoxicity

To quantify cytotoxic effector programs across T cell states, a T cell cytotoxicity module score was computed using Seurat’s AddModuleScore() function on normalized RNA assay data. The gene set included *FGFBP2, CX3CR1, FCGR3A, S1PR5, PLAC8, FGR, C1orf21, SPON2, CD300A, TGFBR3, PLEK, S1PR1, EFHD2, KLRF1, FAM65B, C1orf162, STK38, SORL1, FCRL6, TRDC, EMP3, CCND3, KLRB1, SAMD3, ARL4C, IL7R,* and *GNLY*. Resulting cytotoxicity scores were compared between treatment groups and across time points, as shown in Figure 2H.

#### Subclustering of Tregs

To investigate treatment-associated heterogeneity within regulatory T cells, the integrated reference-mapped T cell dataset was first subset to the predicted Treg population. To ensure analysis of bona fide regulatory T cells, this subset was further restricted to FOXP3-expressing cells prior to reclustering. The resulting FOXP3⁺ Treg population was then reprocessed by normalizing and scaling gene expression values, identifying variable features, and performing principal component analysis. Principal components selected based on variance explained were used to construct neighborhood graphs, define clusters using the Louvain algorithm, and generate UMAP embeddings for visualization. Marker gene expression was examined using Seurat’s FeaturePlot() and Nebulosa’s density representations to evaluate canonical Treg markers (*FOXP3, IL2RA, IKZF2, CTLA4*) alongside cytotoxicity-associated genes (*GZMA, GZMB, GNLY, NKG7*). This refined Treg dataset was used to compare cluster composition across treatment arms and to identify treatment-induced cytotoxic Treg states.

#### TCR V(D)J repertoire analysis

TCR V(D)J contigs generated by Cell Ranger were processed using the scRepertoire package and linked to their corresponding transcriptomic profiles in Seurat. Only cells with productive paired TRA–TRB receptors were retained for analysis. Clonotypes were defined according to scRepertoire conventions, relying on matching TRA and TRB CDR3 amino acid sequences and V/J gene usage. Clonal abundances were calculated for each sample and were classified into established frequency categories: hyperexpanded, large, medium, small, and rare clones.

Longitudinal tracking was carried out by assigning clonotypes detected exclusively before treatment as T_pre and those detected only after treatment as T_post on a per-patient basis. Clonal skewing was quantified using Gini indices, calculated with the ineq R package. One Gini metric described inequality in clonal abundance distributions within pan T cells, CD4⁺ T cells, and CD8⁺ T cells. A second spatial Gini metric quantified inequality in the distribution of specific T cell subsets across UMAP space by discretizing UMAP coordinates into uniform bins and computing the Gini coefficient based on cell density across bins. To avoid sampling bias, all paired analyses were performed on datasets downsampled to the smallest matched T cell count.

#### Single-cell ATAC–seq processing and analysis of Tregs

Isolated Tregs of BTCE–DRd–treated NDMM patients were profiled using the 10x Genomics Chromium Single Cell Multiome ATAC platform, and processed with Cell Ranger ARC (10x Genomics) against the GRCh38/hg38 reference genome to generate fragment files and peak-by-cell count matrices. For each sample and time point (pre- and post-therapy), we imported the filtered peak-by-cell matrix (filtered_feature_bc_matrix.h5) and fragment file (atac_fragments.tsv.gz) into R (v4.4) using Seurat and Signac. Peaks were restricted to standard chromosomes and converted to genomic ranges. Gene annotations were obtained from *EnsDb.Hsapiens.v86*, harmonized to the hg38 genome build, and attached to the chromatin assay. A chromatin assay was created for each sample (Signac CreateChromatinAssay()) using the peak-by-cell count matrix, fragment file, and hg38 annotations, requiring a minimum of 10 cells per peak. Cells were filtered based on standard quality-control metrics, retaining those with moderate ATAC library complexity (approximately 10³–10⁵ fragments per cell) and low mitochondrial content (<5%). Peaks overlapping extremely small (<20 bp) or very large (>10 kb) regions were removed. For each sample, chromatin accessibility was normalized using term frequency–inverse document frequency (TF–IDF) transformation, high-variance features were selected (FindTopFeatures()), and dimensionality reduction was performed using latent semantic indexing (LSI; RunSVD()). UMAP embeddings were computed from LSI components 2–50 to visualize chromatin accessibility states.

To enable joint analysis across patients and time points, we first defined a unified peak set by taking the genomic union of sample-specific peak sets (Signac reduce() on GRanges) and applying the same width filters (20–10,000 bp). For each sample, fragments were re-quantified over this common peak set using FeatureMatrix(), and a new chromatin assay was constructed with the unified peaks. These unified-peak assays were reprocessed independently (TF–IDF, FindTopFeatures, LSI, and UMAP as above). Seurat objects were then merged, and cross-sample integration of chromatin accessibility was performed using Signac’s LSI-based integration workflow. Briefly, we identified integration anchors (FindIntegrationAnchors(), reduction = “rlsi”) and integrated the LSI embeddings across samples (IntegrateEmbeddings()) to obtain a shared low-dimensional representation (“integrated_lsi”), followed by UMAP visualization (RunUMAP()).

Differential chromatin accessibility between pre- and post-therapy Tregs was assessed at the peak level using Signac, contrasting cells from post-therapy samples against the corresponding pre-therapy samples at the patient level. Peaks with significant changes in accessibility were defined as differentially accessible (DA) based on adjusted p-values (Benjamini–Hochberg false discovery rate). Motif enrichment analysis was carried out on DA peak sets using Signac, testing for over-representation of transcription factor binding motifs within DA regions compared to the background set of all unified peaks. Enrichment statistics and fold-enrichment values for AP-1 family motifs (including BATF/JUNB-containing motifs) were used to generate the motif enrichment summaries and plots shown in the figures.

### Cell lines

B-ALL cell line Nalm-6 gfp luc was kindly gifted from the Blaeschke Lab at the German Cancer Research Center. Multiple Myeloma cell line MM1.S gfp luc was obtained from the Max Delbrück Center for Molecular Medicine (MDC). Cells were cultured at 37°C with 5% CO2 in RPMI1640 media (Sigma-Aldrich, Cat# R7388) supplemented with 10% FBS (Gibco, Cat# A5256701), and Penicillin-Streptomycin 1X (Sigma, P4458-100ML).

### Flow cytometry analysis of target markers on cell lines

Cell lines were stained with anti BCMA - APC (Miltenyi, clone: REA315, Cat# 130-131-092), anti GPCR5D - AF647 (R&D Systems, clone: 571932, Cat# FAB63001R-100UG), anti CD19 - BUV496 (BD, clone: SJ25C1, Cat# 612938), and Fixable Viability Dye eFluor™ 780 (eBioscience™, Cat# 65-0865-18) for 30min at 4°C. Subsequently, stained samples were washed and resuspended in FACS buffer and analyzed on the Cytek^®^ Aurora FACS machine. GFP BrightComp eBeads™ (Invitrogen, Cat# A10514) were used for spectral unmixing.

### Co-culture system

To investigate the effects of interactions of regulatory T cells with Multiple Myeloma or Acute lymphoblastic leukemia under different *in vitro* conditions, mono- and co-cultures were used. Purified healthy-donor (HD) CD4^+^ CD127^low^ CD25^+^ regulatory T cells - isolated as described above - were cocultured with Luciferase^+^ target cell lines in a 96-well plate (5×10^4^ and 1×10^4^ cells, respectively) or 384-well plate (1×10^4^ and 2×10^3^ cells, respectively) in RPMI medium with 10% FBS and an E:T ratio of 5:1. Treatment conditions included Teclistamab (Hölzel Diagnostika, Cat# HY-P99392-1mg), Blinatumomab (Hölzel Diagnostika, Cat# A3175-1mg), Talquetamab (Hölzel Diagnostika, Cat# HY-P99394-1mg), Ultra-LEAF™ Purified anti-human CD3 Antibody (Biolegend, #317326) and Lenalidomide (Sigma-Aldrich, #SML2283-100MG) using concentrations as indicated in the respective experiments. As indicated PanT cells were isolated from the same healthy donors using the Pan T Cell Isolation Kit, human (Miltenyi, Cat#130-096-535) and used as controls. Following 48 and 72 hours of coculture, samples were processed as indicated in the respective methods for each experiment.

### Cytotoxicity assay

To investigate the effects of teclistamab under different *in vitro* conditions, mono- and co-cultures of the luciferase^+^ MM.1S myeloma cell line and purified healthy-donor (HD) CD4^+^ CD127^low^ CD25^+^ regulatory T cells were seeded in a 96-well plate (1×10^4^ and 5×10^4^ cells, respectively) or 384-well plate (2×10^3^ and 1×10^4^ cells, respectively) in RPMI medium with 10% FBS and an E:T ratio of 5:1. Cells were then exposed to different concentrations of Teclistamab (Hölzel Diagnostika, Cat# HY-P99392-1mg) or Talquetamab (Hölzel Diagnostika, Cat# HY-P99394-1mg). Equimolar concentrations of CD3 monoclonal antibody (Ultra-LEAF™ Purified anti-human CD3 Antibody, Biolegend, #317326), were used as controls. Additionally fixed concentrations of Teclistamab (100 ng/ml), equimolar concentrations of OKT3 (52,3 ng/ml or Blinatumomab (37,7 ng/ml) as well as Lenalidomide (1uM) and double treatment with Teclistamab and Lenalidomide were tested. After indicated time of incubation, the viability of myeloma cells was quantified on a Spectramax ID3 plate reader using the Bio-Glo™ Luciferase Assay System (Promega, Cat# G7940).

Additionally, healthy donor PBMCs were stained with anti CD3 - BV421 (BD, clone: SK3, Cat# 563789), anti CD4 - BUV395 (BD, clone: SK3, Cat# 566923), anti CD25 - PE (BD, clone: M-A251, Cat# 555432), anti CD127 - AF647 (BD, clone: HIL-7R-M21, Cat# 558598), and Fixable Viability Dye eFluor™ 780 (eBioscience™, Cat# 65-0865-18) and FACS sorted for CD4^+^ CD127^low^ CD25^+^ regulatory T cells. Subsequently cells were co-cultured as described above and treated with Teclistamab (100 ng/ml) and control as described above.

Similarly Luciferase^+^ Nalm-6 cells were cocultured with purified healthy-donor (HD) CD4^+^ CD127^low^ CD25^+^ regulatory T cells and exposed to different concentrations of Blinatumomab (Hölzel Diagnostika, Cat# A3175-1mg). Equimolar concentrations of CD3 monoclonal antibody (Ultra-LEAF™ Purified anti-human CD3 Antibody, Biolegend, #317326), were used as controls. After indicated time of incubation, the viability of ALL cells was quantified as described above.

### Analysis of cell culture supernatants by Enzyme-Linked Immunosorbent Assay (ELISA)

For the analysis of cell culture supernatants, CD4^+^ CD127^low^ CD25^+^ regulatory T cells were isolated from healthy donors and co-cultured with MM.1S myeloma cells (1×10^6^ cells/ml and 2×10^5^cells/ml, respectively) for 72 hours with either Teclistamab (100 ng/ml), equimolar concentrations of OKT3 (52,3 ng/ml) or Blinatumomab (37,7 ng/ml) as well as Lenalidomide (1uM) and double treatment with Teclitamab and Lenalidomide. Supernatants were harvested and stored at -80°C until analysis. Cytokine concentrations of Granzyme A (Invitrogen, Cat# BMS2232), Granzyme B (Invitrogen, Cat# BMS2027-2), Granulysin (R&D Systems, Cat# DY3138), CCL5 (R&D Systems, Cat# DY278), IL35 (Invitrogen, Cat# EEL067), and IL10 (Invitrogen, Cat# EHIL10) were determined by ELISAs following manufactures guidelines.

### Flow cytometry analysis of intracellular cytokines following Treg coculture

For the characterization of healthy donor CD4^+^ CD127^low^ CD25^+^ regulatory T cells following coculture with MM.1S myeloma cells and treatment with either teclistamab (50 ng/ml), equimolar concentrations of OKT3 (26,15 ng/ml) as well as lenalidomide (1uM) and double treatment with teclistamab and lenalidomide, an intracellular spectral flow cytometry panel was used. At 66hr of culture BD GolgiPlug™ Protein Transport Inhibitor (Containing Brefeldin A) (BD, #555029) was added for 6hr to the coculture to induce intracellular cytokine accumulation within the golgi vesicles. Samples were washed in PBS and subsequently stained. Following live dead staining with Fixable Viability Dye eFluor™ 780 (eBioscience™, Cat# 65-0865-18) for 15min at 4°C surface markers (CD3 - BV786 (BD, clone: SK7, Cat# 563800), CD4 - BUV395 (BD, clone: SK3, Cat# 563550), CD8 - BV605 (BioLegend, clone: SK1, Cat# 344742), CD25 - BV650 (BD, clone: M-A251, Cat# 563719), CD127 - AF647 (BD, clone: HIL-7R-M21, Cat# 558598)) were stained for 30min at 4°C. Cells were washed and permeabilized with the eBioscience™ Foxp3 / Transcription Factor Staining Buffer Set (Invitrogen™, #00-5523-00) following manufactures protocol. Intracellular cytokines (Granzyme A - R718 (BD, clone: CB9, Cat# 566976), Granzyme B - RB780 (BD, clone: GB11, Cat# 568705), Granulysin - AF488 (BD, clone: RB1, Cat# 558254), CCL5 - BV421 (BD, clone: 2D5, Cat# 564754), IFNy - RB613 (BD, clone: B27, Cat# 571095)) were stained together with anti-FoxP3 - PE (BD, clone: 236A/E7, Cat# 560852) antibody for 30min at room temperature in the dark. Following staining samples were washed and resuspended in FACS buffer and analyzed on the Cytek^®^ Aurora. GFP BrightComp eBeads™ (Invitrogen, Cat# A10514) were used for spectral unmixing.

### qPCR

Tregs were collected after 72h coculture with MM1.S cells and washed once with medium to remove residual tumor cells. Cells were counted, pelleted, and RNA was isolated using the RNeasy Micro Kit (Qiagen Cat# 74004) following the manufacturer’s instructions. Subsequently, cDNA was generated from purified RNA using the High-Capacity cDNA Reverse Transcription Kit (Thermofisher Cat# 4374966) by performing a standard reverse-transcription reaction (20 µL total volume) with a 2× RT master mix according to the kit protocol. Quantitative real-time PCR was performed on a QuantStudio™ 3 Real-Time PCR System using 96-well plates, TaqMan™ Fast Advanced Master Mix (ThermoFisher Cat# 4444557), and standard cycling conditions. TaqMan® Gene Expression Assays (ThermoFisher) with customized probes targeting signaling, transcriptional, and effector genes were used, including: (*MAP3K1* (Hs00394890_m1), *MAPK8/JNK1* (Hs01548508_m1), *MAPK14/p38* (Hs01051152_m1), *STAT1* (Hs01013996_m1), *STAT3* (Hs00374280_m1), *STAT5B* (Hs00560026_m1), *TNFAIP3* (Hs00234713_m1), *DUSP4* (Hs01027785_m1), *DUSP16* (Hs00411837_m1), *NFATC1* (Hs00542675_m1), *BATF* (Hs00232390_m1), *JUN* (Hs01103582_s1), *JUNB* (Hs00357891_s1), *FOS* (Hs04194186_s1), *IRF4*(Hs00180031_m1), *PRDM1/BLIMP1* (Hs00153357_m1), *GZMA* (Hs00989184_m1), *GZMB* (Hs00188051_m1), *NKG7* (Hs01120688_g1), *PRF1* (Hs00169473_m1), *IFNG*(Hs00989291_m1), *TOX* (Hs01049519_m1), *CCR8* (Hs00174764_m1), *IKZF2/Helios* (Hs00915979_m1), *IL2RA/CD25* (Hs00907777_m1), *FOXP3* (Hs01085834_m1), *CTLA4* (Hs00175480_m1), *TIGIT* (Hs00545087_m1), *18S rRNA* (Hs99999901_s1), *GAPDH* (Hs99999905_m1), *HPRT1* (Hs99999909_m1), *GUSB* (Hs99999908_m1)). Each condition was measured across three biological replicates, and all qPCR reactions were run in technical duplicates. Expression was normalized to housekeeping genes. Relative gene expression was calculated using the ΔΔCt method.

### Bulk RNA sequencing

RNA of assayed primary human Tregs was isolated using the RNeasy Micro Kit (Qiagen Cat# 74004) according to manufacturer’s instructions. Following RNA isolation, sequencing libraries were generated using the SMARTer Ultra Low RNA Kit (634948, Takara) and sequenced by paired-end sequencing (100 bp) on a NovaSeq X 10B flow cell. The resulting FASTQ files were processed using the Roddy-based workflow RNAseqWorkflow (DKFZ-ODCF, Heidelberg) for alignment, quality control and quantification. Briefly, reads were aligned in two-pass mode using STAR to the reference genome with GENCODE annotations. Duplicate marking was performed with sambamba, and mapping quality was assessed via RNASeQC together with flagstat summaries. Gene-level read counting was carried out using featureCounts (Subread) with paired-end mode and quality threshold set to 255, and strand-unspecific counting applied. Expression values (RPKM/TPM) were computed with a custom script excluding genes from chromosomes X, Y, mitochondrial genome, rRNA and tRNA in the library size estimation. Fusion detection was enabled using Arriba. All steps were run with default parameters unless otherwise stated, and versions of each tool and index (e.g., GENCODE v19) are available on GitHub (https://github.com/DKFZ-ODCF/RNAseqWorkflow). Differential gene expression analysis was performed using DESeq2 (v1.48.2). Gene-level read counts from featureCountsoutput files were imported, aggregated by gene ID, merged across samples, and missing values were set to zero. Sample metadata (donor and treatment) was inferred from sample names, and a DESeq2 dataset was generated using the design formula ∼ donor + treatment to account for donor-specific variation. Genes with low counts (fewer than 10 reads in at least 5 samples) were removed prior to model fitting. Variance-stabilized normalization, size-factor estimation, dispersion modeling, and negative binomial fitting were performed with DESeq(), and differential expression was assessed using contrasts of interest. Resulting log2 fold changes and adjusted p-values were extracted with results(), and downstream visualization (heatmaps) was performed using the pheatmap package.

### Profiling of Treg cell metabolism

Isolated CD4⁺CD127^lowCD25⁺ regulatory T cells (Tregs) or pan T cells were cocultured with MM1.S myeloma cells for 72 hours in the presence of teclistamab (100 ng/mL), as described above. After coculture, CD138⁺ MM1.S cells were depleted using CD138 MicroBeads (Miltenyi Biotec; Cat# 130-051-301) according to the manufacturer’s instructions. The remaining Tregs and pan T cells were then subjected to metabolic profiling using the XF T Cell Persistence Assay on a Seahorse XFe96 Analyzer (Agilent; Cat# 103772-100).

### Quantification and statistical analysis

Data are represented as individual values or as mean ± SEM, as indicated. Group sizes (n) and applied statistical tests are indicated in figure legends. Statistical significance and multiple hypothesis testing corrections were assessed as indicated in figure legends. All reported p values are two-tailed. All analyses were performed using either R v4.3.2 (www.R-project.org) or GraphPad Prism 10.0. For functional experiments, bone marrow samples were blinded to the experiment performers. Shannon and all others indices were calculated using diversity() vegan R package or Gini() as stated in methods above.

Due to the nature of this study, sample size determination was not applicable, as all available samples were included in this study. All cells passing QC (**Figure S1-2**, methods) were included in downstream analyses on a single-cell basis.

### Data visualization

Tabular data from single-cell sequencing analyses above were processed using the tidyverse suite of packages [https://CRAN.R-project.org/package=tidyverse] and visualized in the R programming environment using the ggplot2 package or the Python programming environment using the matplotlib package. Data from all other analyses were visualized using GraphPad Prism 10.0. Figures were produced using Adobe Illustrator 2024.

## Data Availability

The data generated in this study are available via Mendeley Data. The repository will be made public upon publication of the peer-reviewed manuscript.

## Data and code availability

The data generated in this study are available via Mendeley Data. A private preview link has been provided to the editors and reviewers for peer review. The repository will be made public upon publication of the peer-reviewed manuscript.

## Acknowledgements

We thank all members of the Friedrich laboratory for their support and useful discussions. We are grateful to the High Throughput Sequencing Unit of the Genomics and Proteomics Core Facility, and the Omics IT and Data Management Core Facility for their technical support. We thank the Clinical Myeloma Registry and Biobank at Heidelberg University Hospital, funded by the Dietmar Hopp Stiftung and the Paula and Roger Riney Foundation. This study includes samples provided by the NCT Cell and Liquid Biobank (NCT CLB), a member of the Biomaterial Bank Heidelberg (BMBH). We thank the GMMG (German Speaking Myeloma Multicenter Group) for providing the samples used in this study.

This study was supported by the Deutsche José Carreras Leukämie-Stiftung (DJCLS, PI: M.J. Friedrich; Project-ID DJCLS 01ZI/2022); the Dr. Rolf M. Schwiete Stiftung (PI: M.J. Friedrich; Project-ID 2025-018); the Else Kröner-Fresenius-Stiftung (PI: M.J. Friedrich; Project-ID 2025_EKMS.52); the Stiftung Deutsche Krebshilfe (PI: M.J. Friedrich; Project-ID 70117225); the Dietmar Hopp Foundation (PI: M.J. and A.T.) and the Deutsche Forschungsgemeinschaft (DFG, German Research Foundation) – SFB 1709/1 2025 – 533056198.

## Author information

### Contributions

J.M., J.H.F. and M.J.F. conceived the project. J.M. and A.D.M. designed all experiments with input from M.J.F. and other authors. J.M. and A.D.M. performed and analyzed all *in vitro* experiments.

A.S. performed spectral flow cytometry experiments and analyses. J.M., J.H.F., A.D.M. and V.T. performed longitudinal single-cell RNA/TCR/ATAC-seq and bulk RNA-seq experiments. J.M. and N.K. performed longitudinal single-cell RNA/TCR/ATAC-seq and bulk RNA-seq analyses. T.R.W., R.O., and A.K. provided assistance with *in vitro* and sequencing experiments. J.Z., S.C. and C.F. provided critical mentorship and guidance in clinical analyses. S.H. and A.B. processed and analyzed primary human bone marrow and blood specimens. M.J.F. supervised this research and experimental design with support from A.T., M.H. C.M.T., H.G., H.E., L.R., M.S.R. and N.W. J.M., J.H.F. and M.J.F. wrote the manuscript with input from all authors.

