## Supplementary Materials for "CD3-engaging bispecific antibodies convert human regulatory T cells into cytotoxic effectors"

**Disclaimer:** This article is a preprint reporting new medical research and has not been peer-reviewed; it should not be used to guide clinical practice or be reported in the press as conclusive

### Supplementary Materials for

### CD3-engaging bispecific antibodies convert human regulatory T cells into cytotoxic effectors

Julius J. Michel, Jan H. Frenking, Anna D. Metzler, Antonia Schach, Erin W. Meermeier, Vera Thiel, Niklas Kehl, Tim R. Wagner, Julian Zoller, Sven Cuntz, Cornelius Funk, René Onken, Alanna Kirschner, Stefanie Huhn, Alexander Brobeil, Andreas Trumpp, Mohammad Rahbari, Mathias Heikenwälder, Carsten Müller-Tidow, P. Leif Bergsagel, Hartmut Goldschmidt, Hermann Einsele, Leo Rasche, Marta Chesi, Marc S. Raab, Niels Weinhold, and Mirco J. Friedrich.

**The PDF file includes:**

Supplementary Figures S1-S8

Supplementary Table 1

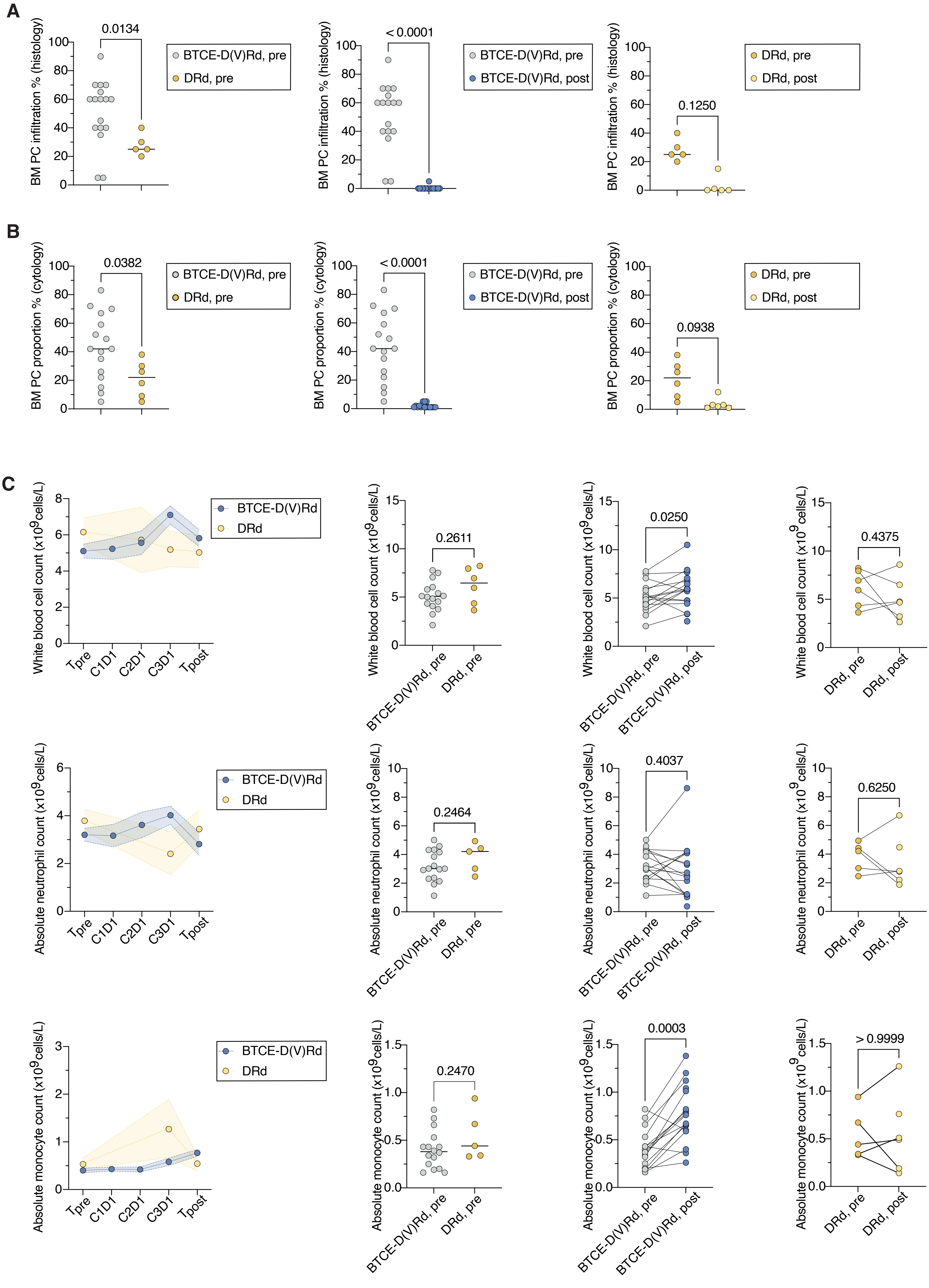
A
BM PC infiltration % (histology)
0.0134
BTCE-D(V)Rd, pre
DRd, pre
< 0.0001
BTCE-D(V)Rd, post
0.1250
DRd, post
B
BM PC proportion % (cytology)
0.0382
< 0.0001
0.0938
C
White blood cell count (x10⁹cells/L)
Tpre
C1D1
C2D1
C3D1
Tpost
BTCE-D(V)Rd
DRd
0.2611
0.0250
0.4375
Absolute neutrophil count (x10⁹cells/L)
0.2464
0.4037
0.6250
Absolute monocyte count (x10⁹cells/L)
0.2470
0.0003
> 0.9999

**Supplementary Figure S1. Front-line BTCE therapy reduces bone marrow plasma cell burden and preserves peripheral leukocyte counts in NDMM patients.**

**(A)** Quantification of bone marrow plasma cell (PC) infiltration (percentage by histology) in patients with newly-diagnosed multiple myeloma (NDMM) from the BTCE-D(V)Rd group (grey and blue) and the DRd group (yellow) at pre-treatment (pre / before C1) and post-treatment (post / after C3) time points. P-values are indicated and were calculated by Mann Whitney test or Wilcoxon matched-pairs signed rank test. BTCE, bispecific T-cell engager. DRD, daratumumab + lenalidomide + dexamethasone. D-VRd, daratumumab + bortezomib + lenalidomide + dexamethasone. BTCE-D(V)Rd, pre (n=17); BTCE-D(V)Rd, post (n=16); DRd, pre (n=5); DRd, post (n=5).

**(B)** Quantification of bone marrow plasma cells (percentage by cytology) in patients with NDMM in the BTCE-D(V)Rd group (grey and blue) and the DRd group (yellow) at pre (before C1) and post (after C3). P-values are indicated and were calculated as described above. BTCE-D(V)Rd, pre (n=16); BTCE-D(V)Rd, post (n=16); DRd, pre (n=6); DRd, post (n=6).

**(C)** Line and dot plots showing changes in peripheral blood White Blood Cell (WBC) count, Absolute Neutrophil Count (ANC), and Absolute Monocyte Count (AMC) over the course of treatment in the BTCE group (grey and blue) and CTR group (yellow). Paired dot plots on the right compare pre- (before C1) versus post- (after C3) treatment values within each group, with p-values indicated and calculated as described above. BTCE-D(V)Rd, pre (n=16); BTCE-D(V)Rd, post (n=17); DRd, pre (n=5-6); DRd, post (n=6).

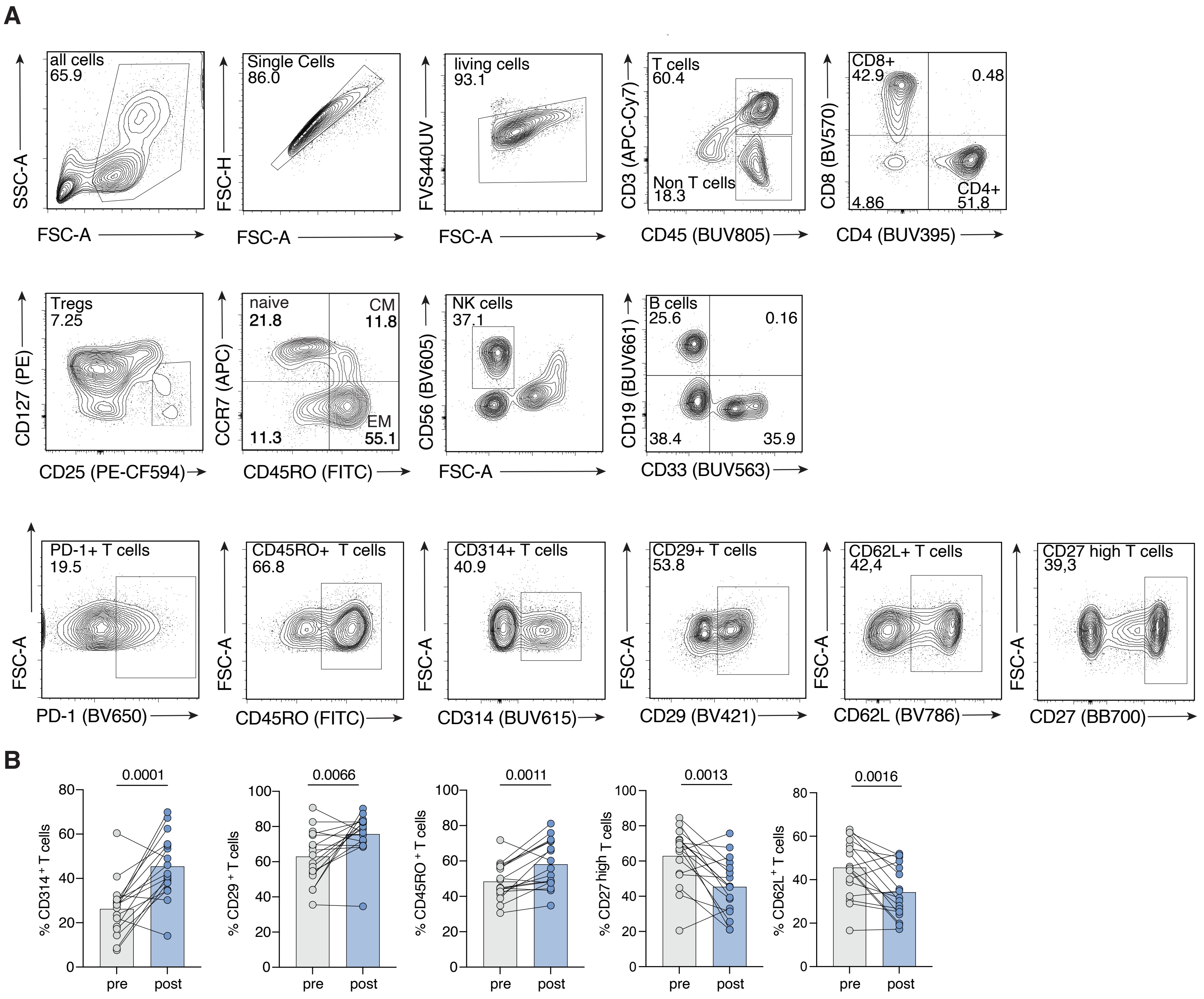
A
all cells
65.9
SSC-A
FSC-A
Single Cells
86.0
FSC-H
FSC-A
living cells
93.1
FVS440UV
FSC-A
T cells
60.4
Non T cells
18.3
CD3 (APC-Cy7)
CD45 (BUV805)
CD8+
42.9
0.48
4.86
CD4+
51.8
CD8 (BV570)
CD4 (BUV395)
Tregs
7.25
CD127 (PE)
CD25 (PE-CF594)
naive
21.8
CM
11.8
EM
11.3
55.1
CCR7 (APC)
CD45RO (FITC)
NK cells
37.1
CD56 (BV605)
FSC-A
B cells
25.6
0.16
38.4
35.9
CD19 (BUV661)
CD33 (BUV563)
PD-1+ T cells
19.5
FSC-A
PD-1 (BV650)
CD45RO+ T cells
66.8
FSC-A
CD45RO (FITC)
CD314+ T cells
40.9
FSC-A
CD314 (BUV615)
CD29+ T cells
53.8
FSC-A
CD29 (BV421)
CD62L+ T cells
42,4
FSC-A
CD62L (BV786)
CD27 high T cells
39,3
FSC-A
CD27 (BB700)
B
0.0001
% CD314 + T cells
pre
post
0.0066
% CD29 + T cells
pre
post
0.0011
% CD45RO + T cells
pre
post
0.0013
% CD27 high T cells
pre
post
0.0016
% CD62L + T cells
pre
post

**Supplementary Figure S2. Flow cytometric profiling of peripheral T cell subsets and activation states before and after therapy.**

**(A)** Representative flow cytometric gating used to identify major T cell subsets from peripheral blood. Sequential gates illustrate the selection of single cells (FSC-A/FSC-H), live cells (FVS440UV$^{-}$), T cells (CD3$^{+}$CD45$^{+}$), CD8$^{+}$ and CD4$^{+}$ T cell populations, Tregs (CD3$^{+}$CD4$^{+}$ CD25$^{+}$CD127$^{low}$), NK cells (CD3$^{-}$CD56$^{+}$), B cells(CD3$^{-}$CD19$^{+}$), followed by delineation of T cell memory subsets, naïve, central memory (CM), and effector memory (EM), based on CCR7 and CD45RO expression. Expression of markers associated with cytotoxic activation/function (PD-1, NKG7/CD314, CD29), memory differentiation (CD45RO), and less differentiated phenotypes (CD62L, CD27^high) are also shown.

**(B)** Flow cytometric quantification of CD3$^{+}$ T cells expressing markers of cytotoxic activation/function (NKG7/CD314, CD29), memory differentiation (CD45RO), and less differentiated phenotypes (CD62L, CD27$^{high}$) in peripheral blood before and after treatment. P values were calculated using paired t tests for each comparison.

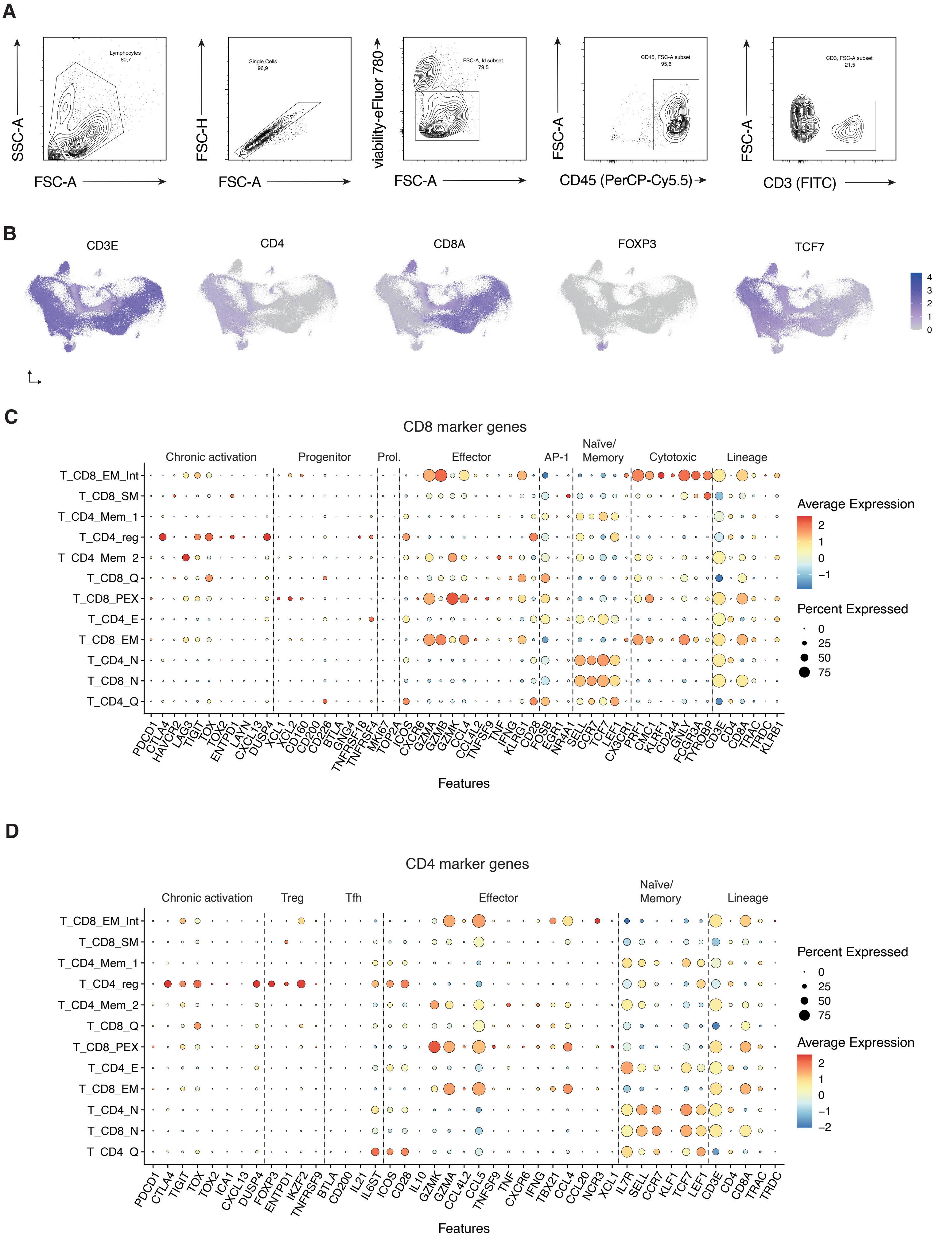
A
SSC-A
FSC-A
Lymphocytes
80,7
FSC-H
FSC-A
Single Cells
96,9
viability-eFluor 780
FSC-A
FSC-A, ld subset
79,5
FSC-A
CD45 (PerCP-Cy5.5)
CD45, FSC-A subset
95,6
FSC-A
CD3 (FITC)
CD3, FSC-A subset
21,5
B
CD3E
CD4
CD8A
FOXP3
TCF7
4
3
2
1
0
C
CD8 marker genes
Chronic activation
Progenitor
Prol.
Effector
AP-1
Naïve/
Memory
Cytotoxic
Lineage
T_CD8_EM_Int
T_CD8_SM
T_CD4_Mem_1
T_CD4_reg
T_CD4_Mem_2
T_CD8_Q
T_CD8_PEX
T_CD4_E
T_CD8_EM
T_CD4_N
T_CD8_N
T_CD4_Q
PDCD1
CTLA4
HAVCR2
LAG3
TIGIT
TOX
TOX2
ENTPD1
LAYN
CXCL13
DUSP4
XCL1
XCL2
CD160
CD200
CD226
BTLA
GNG4
TNFRSF18
TNFRSF4
MKI67
TOP2A
ICOS
CXCR6
GZMA
GZMB
GZMK
CCL4
CCL4L2
TNFSF9
TNF
IFNG
KLRG1
CD28
FOSB
EGR1
NR4A1
SELL
CCR7
TCF7
LEF1
CX3CR1
PRF1
CMC1
KLRF1
CD244
GNLY
FCGR3A
TYROBP
CD3E
CD4
CD8A
TRAC
TRDC
KLRB1
Features
Average Expression
2
1
0
−1
Percent Expressed
0
25
50
75
D
CD4 marker genes
Chronic activation
Treg
Tfh
Effector
Naïve/
Memory
Lineage
T_CD8_EM_Int
T_CD8_SM
T_CD4_Mem_1
T_CD4_reg
T_CD4_Mem_2
T_CD8_Q
T_CD8_PEX
T_CD4_E
T_CD8_EM
T_CD4_N
T_CD8_N
T_CD4_Q
PDCD1
CTLA4
TIGIT
TOX
TOX2
ICA1
CXCL13
DUSP4
FOXP3
ENTPD1
IKZF2
TNFRSF9
BTLA
CD200
IL21
IL6ST
ICOS
CD28
IL10
GZMK
GZMA
CCL4L2
CCL5
TNFSF9
TNF
CXCR6
IFNG
TBX21
CCL4
CCL20
NCR3
XCL1
IL7R
SELL
CCR7
KLF1
TCF7
LEF1
CD3E
CD4
CD8A
TRAC
TRDC
Features
Percent Expressed
0
25
50
75
Average Expression
2
1
0
−1
−2

**Supplementary Figure S3. Single-cell transcriptomic and TCR profiling of bone marrow T cells following Teclistamab-DRd or DRd therapy.**

**(A)** Representative flow cytometric gating used to isolate $CD45^{+}CD3^{+}$ T cells from bone marrow aspirates. Sorted populations were processed for single-cell 5′ RNA-seq and paired TCR V(D)J profiling using the 10x Genomics Chromium platform.

**(B)** UMAP embedding of bone marrow T cells from patients treated with teclistamab-DRd or DRd (control). UMAP visualization of T cells from all included patients, with overlaid single-cell RNA expression used to annotate canonical T cell populations.

**(C–D)** Canonical marker gene expression across T cell subsets. Dot plots showing average expression and detection frequency of curated canonical marker genes across $CD8^{+}$ **(C)** and $CD4^{+}$ **(D)** T cell clusters, based on established gene signatures.

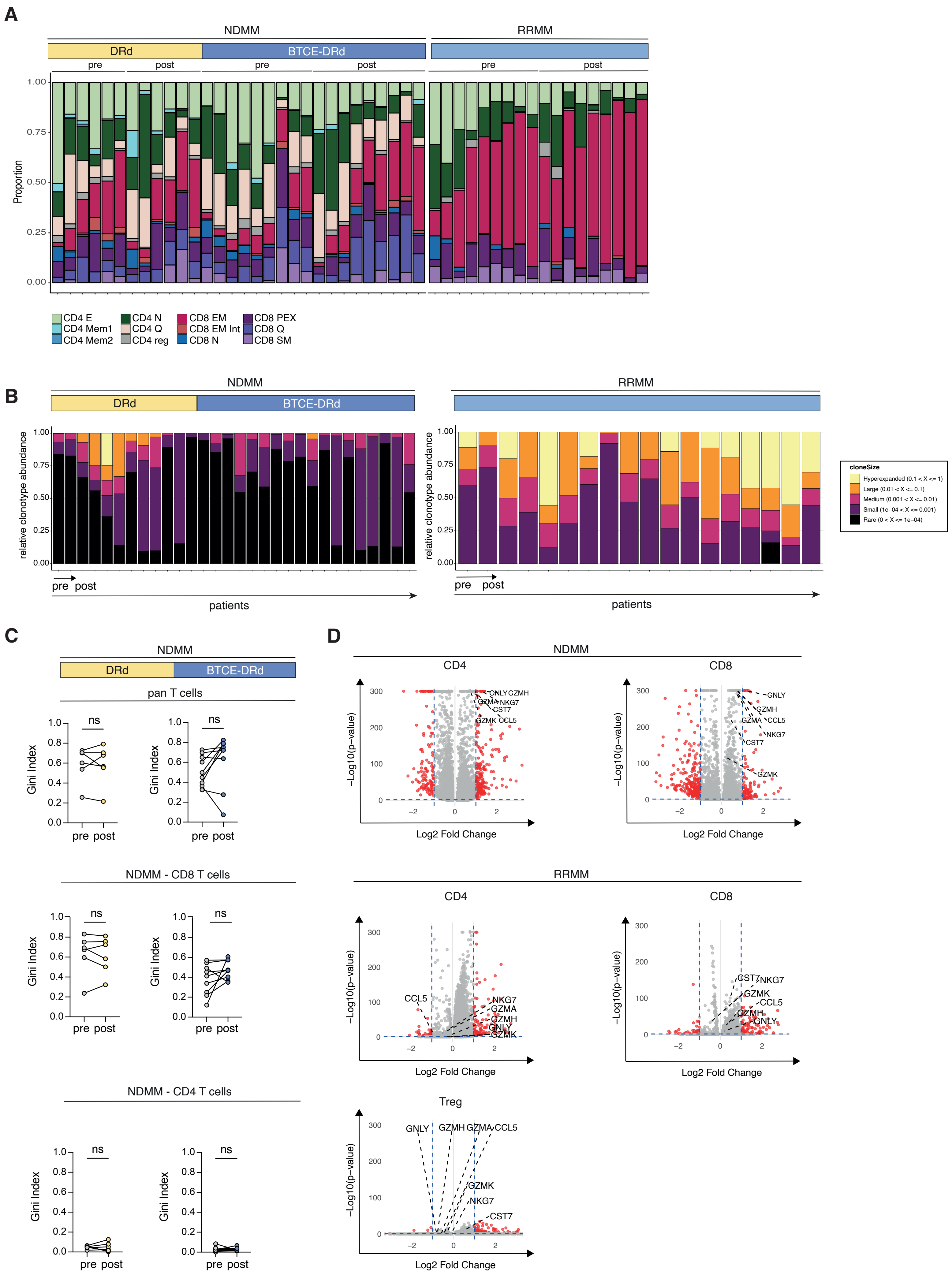

A
NDMM
RRMM
DRd
BTCE-DRd
pre
post
Proportion
1.00
0.75
0.50
0.25
0.00
CD4 E
CD4 Mem1
CD4 Mem2
CD4 N
CD4 Q
CD4 reg
CD8 EM
CD8 EM Int
CD8 N
CD8 PEX
CD8 Q
CD8 SM
B
relative clonotype abundance
pre post
patients
cloneSize
Hyperexpanded (0.1 < X <= 1)
Large (0.01 < X <= 0.1)
Medium (0.001 < X <= 0.01)
Small (1e−04 < X <= 0.001)
Rare (0 < X <= 1e−04)
C
pan T cells
NDMM - CD8 T cells
NDMM - CD4 T cells
Gini Index
ns
D
CD4
CD8
Treg
−Log10(p−value)
Log2 Fold Change
GNLY
GZMH
GZMA
NKG7
CST7
GZMK
CCL5

**Supplementary Figure S4. T cell cluster composition, clonal architecture, and transcriptional remodeling across NDMM and RRMM cohorts.**

**(A)** T-cell cluster composition across patients with newly-diagnosed multiple myeloma (NDMM) and patients with RRMM (relapsed/refractory multiple myeloma). Bar plots showing the distribution of transcriptionally defined T-cell clusters in all patients with NDMM or RRMM, including both pre- (before C1) and post- (after C3) therapy samples. Each bar represents a single sample.

**(B)** Clonotype abundance categories across patient samples. Relative clonotype abundance is shown for each sample (with pre- and post-therapy samples from the same patient displayed consecutively). Clonotypes are grouped into abundance classes: hyperexpanded (0.1–1.0), large (0.01–0.1), medium (0.001–0.01), small (0.0001–0.001), and rare (<0.0001).

**(C)** Gini indices quantifying T cell clonal distribution. Gini indices are shown for pan T cells, $CD4^+$ T cells, and $CD8^+$ T cells, comparing BTCE-DRd and DRd groups. All samples were down-sampled to the same minimum paired T cell count, as in the main figures. BTCE, bispecific T-cell engager. DRd, daratumumab + lenalidomide + dexamethasone.

**(D)** Differential gene expression in $CD4^+$ and $CD8^+$ T cell subsets. Volcano plots showing differential gene expression between post-therapy and pre-therapy T cells in NDMM patients for $CD4^+$ and $CD8^+$ subsets (left panels), and corresponding analyses for RRMM patients in $CD4^+$, $CD8^+$, and Treg subsets (right panels).

A

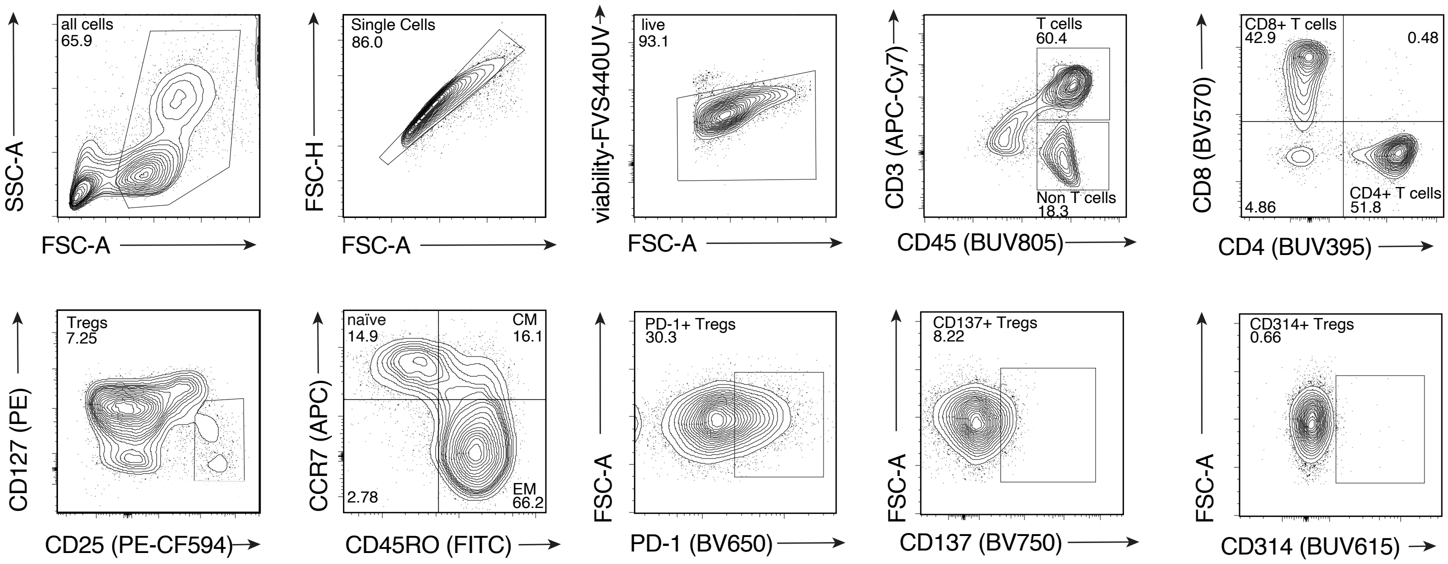
all cells
65.9
SSC-A
FSC-A
Single Cells
86.0
FSC-H
FSC-A
live
93.1
viability-FVS440UV
FSC-A
T cells
60.4
Non T cells
18.3
CD3 (APC-Cy7)
CD45 (BUV805)
CD8+ T cells
42.9
0.48
4.86
CD4+ T cells
51.8
CD8 (BV570)
CD4 (BUV395)
Tregs
7.25
CD127 (PE)
CD25 (PE-CF594)
naïve
14.9
CM
16.1
2.78
EM
66.2
CCR7 (APC)
CD45RO (FITC)
PD-1+ Tregs
30.3
FSC-A
PD-1 (BV650)
CD137+ Tregs
8.22
FSC-A
CD137 (BV750)
CD314+ Tregs
0.66
FSC-A
CD314 (BUV615)

B

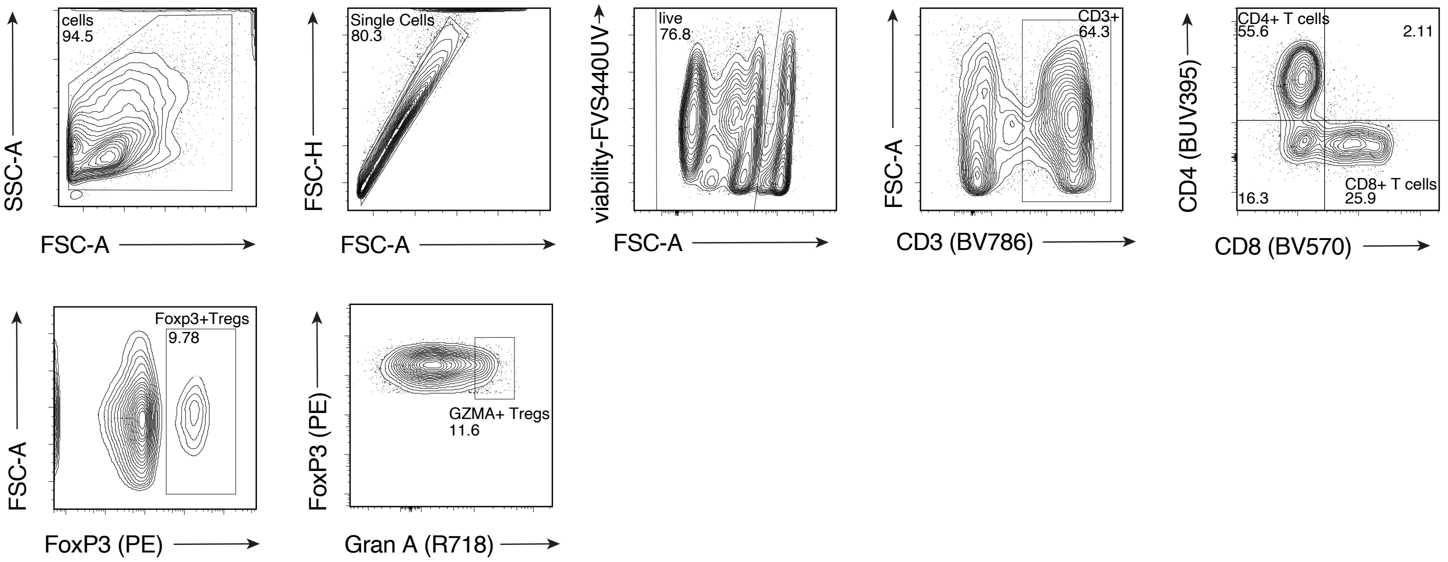
cells
94.5
SSC-A
FSC-A
Single Cells
80.3
FSC-H
FSC-A
live
76.8
viability-FVS440UV
FSC-A
CD3+
64.3
FSC-A
CD3 (BV786)
CD4+ T cells
55.6
2.11
16.3
CD8+ T cells
25.9
CD4 (BUV395)
CD8 (BV570)
Foxp3+Tregs
9.78
FSC-A
FoxP3 (PE)
GZMA+ Tregs
11.6
FoxP3 (PE)
Gran A (R718)

C

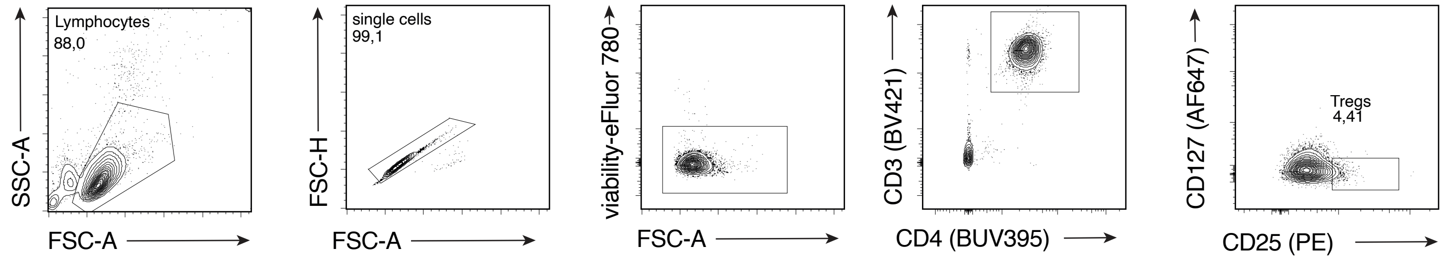
Lymphocytes
88,0
SSC-A
FSC-A
single cells
99,1
FSC-H
FSC-A
viability-eFluor 780
FSC-A
CD3 (BV421)
CD4 (BUV395)
Tregs
4,41
CD127 (AF647)
CD25 (PE)

**Supplementary Figure S5. Flow cytometric identification of major immune cell populations and regulatory T cells in patient PBMCs.**

**(A)** Representative flow cytometric gating used to identify major immune cell populations from patient peripheral blood mononuclear cells (PBMCs).

**(B)** Representative intracellular flow cytometry workflow used to identify $FOXP3^{+}$ regulatory T cells and assess intracellular marker expression within the Treg compartment of patient PBMCs.

**(C)** Representative flow cytometric gating used to isolate $CD3^{+}CD4^{+}CD127^{low}CD25^{+}$ regulatory T cells from healthy donor PBMCs.

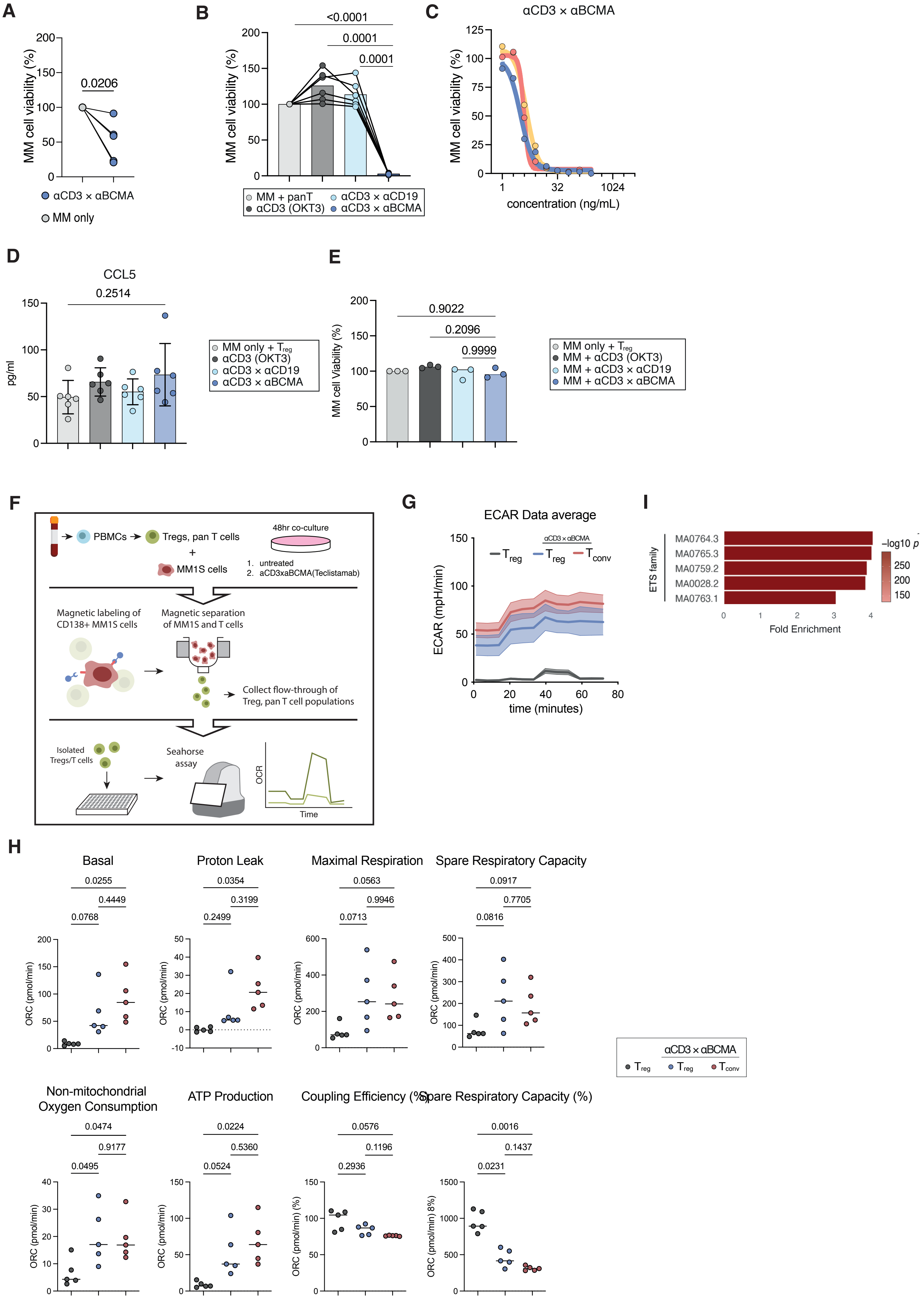

A
B
C
D
E
F
G
H
I
MM cell viability (%)
0.0206
αCD3 × αBCMA
MM only
<0.0001
0.0001
0.0001
MM + panT
αCD3 (OKT3)
αCD3 × αCD19
αCD3 × αBCMA
concentration (ng/mL)
CCL5
0.2514
pg/ml
MM only + Treg
0.9022
0.2096
0.9999
MM cell Viability (%)
MM + αCD3 (OKT3)
MM + αCD3 × αCD19
MM + αCD3 × αBCMA
PBMCs
Tregs, pan T cells
48hr co-culture
MM1S cells
1. untreated
2. aCD3xaBCMA(Teclistamab)
Magnetic labeling of CD138+ MM1S cells
Magnetic separation of MM1S and T cells
Collect flow-through of Treg, pan T cell populations
Isolated Tregs/T cells
Seahorse assay
OCR
Time
ECAR Data average
ECAR (mpH/min)
time (minutes)
Treg
Tconv
ETS family
MA0764.3
MA0765.3
MA0759.2
MA0028.2
MA0763.1
Fold Enrichment
−log10 p
Basal
0.0255
0.4449
0.0768
Proton Leak
0.0354
0.3199
0.2499
Maximal Respiration
0.0563
0.9946
0.0713
Spare Respiratory Capacity
0.0917
0.7705
0.0816
ORC (pmol/min)
Non-mitochondrial Oxygen Consumption
0.0474
0.9177
0.0495
ATP Production
0.0224
0.5360
0.0524
Coupling Efficiency (%)
0.0576
0.1196
0.2936
Spare Respiratory Capacity (%)
0.0016
0.1437
0.0231
ORC (pmol/min) (%)
ORC (pmol/min) 8%)

**Supplementary Figure S6. Functional and metabolic characterization of regulatory T cell and pan-T cell responses to bispecific antibody stimulation.**

**(A)** Sorted regulatory T cells were cocultured with MM1.S myeloma cells and treated with 100 ng/mL Teclistamab for 48 h; MM1.S viability was quantified via luciferase-based luminescence.

**(B)** Viability of MM1.S cells following 48-h coculture with healthy donor pan T cells treated with Blinatumomab (αCD3×αCD19, 37.7 ng/mL), OKT3 (αCD3, 52.3 ng/mL), or Teclistamab (αCD3×αBCMA, 100 ng/mL). Statistical significance was assessed by two-way ANOVA with multiple comparisons (n = 6).

**(C)** Viability of MM1.S cells following 48-h coculture with healthy donor pan T cells treated with increasing concentrations of Teclistamab (n = 3).

**(D)** Quantification of CCL5 secretion during 72-h coculture of MM1.S cells with healthy donor regulatory T cells treated with equimolar concentrations of Blinatumomab (37.7 ng/mL), OKT3 (52.3 ng/mL), or Teclistamab (100 ng/mL). Statistical significance was determined by two-way ANOVA (n = 6).

**(E)** Viability of MM1.S cells directly treated for 48 h with Blinatumomab (37.7 ng/mL), OKT3 (52.3 ng/mL), or Teclistamab (100 ng/mL), in the absence of T cells. Two-way ANOVA with multiple comparisons (n = 3).

**(F)** Schematic workflow of the coculture system and Seahorse metabolic assay used for experiments in Figure 3P and S6H.

**(G)** Extracellular acidification rate (ECAR) of untreated and Teclistamab-treated (100 ng/mL) regulatory T cells and pan T cells after 48-h coculture, measured using the Seahorse XF T Cell Metabolic Profiling Kit. Statistical significance was assessed by repeated-measures one-way ANOVA (n = 5).

**(H)** Oxygen consumption rates (OCR) in pmol/min for basal respiration, proton leak, maximal respiration, spare respiratory capacity, non-mitochondrial oxygen consumption, ATP production, coupling efficiency (%), and spare respiratory capacity quantified during XF Cell Mito Stress Test. Repeated-measures one-way ANOVA (n=5).

**(I)** ETS motif enrichment analysis of differentially accessible regions, showing fold enrichment of representative ETS family motifs.

A

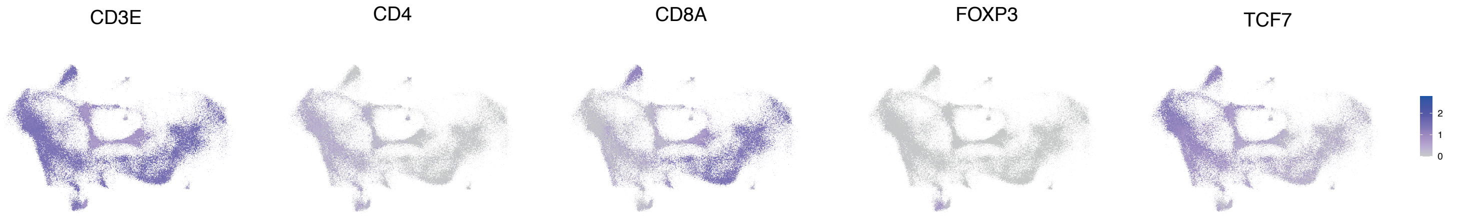
CD3E
CD4
CD8A
FOXP3
TCF7
2
1
0

B

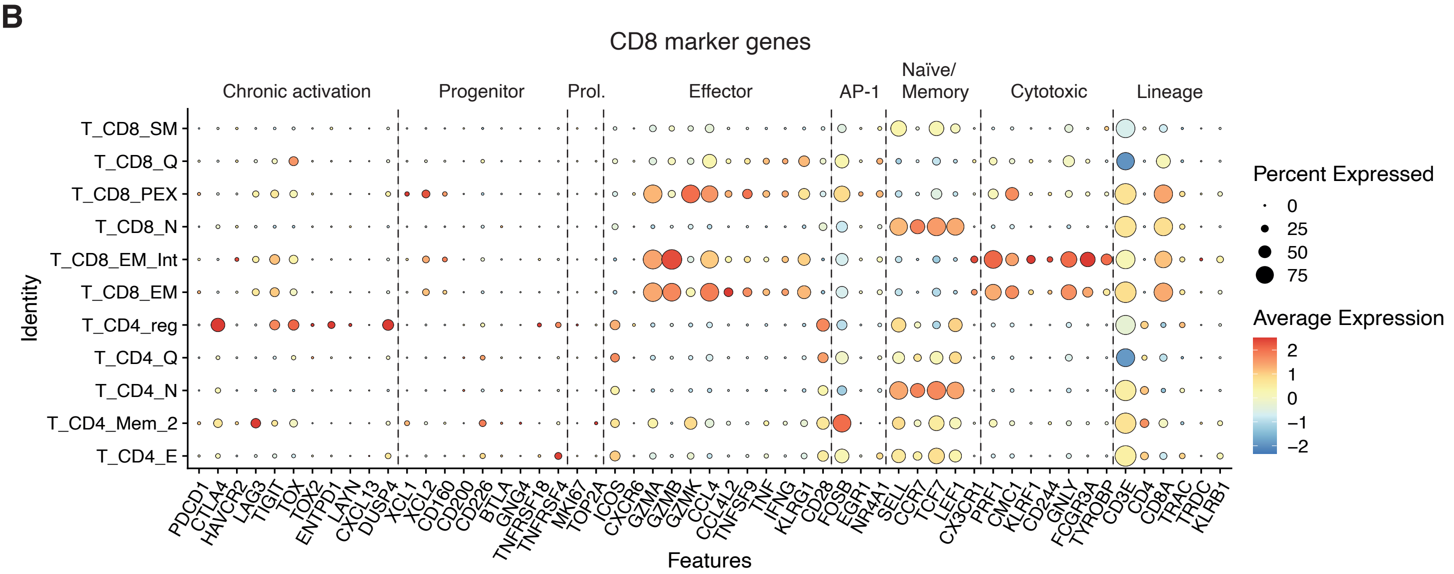
CD8 marker genes
Chronic activation
Progenitor
Prol.
Effector
AP-1
Naïve/
Memory
Cytotoxic
Lineage
T_CD8_SM
T_CD8_Q
T_CD8_PEX
T_CD8_N
T_CD8_EM_Int
T_CD8_EM
T_CD4_reg
T_CD4_Q
T_CD4_N
T_CD4_Mem_2
T_CD4_E
Identity
PDCD1 CTLA4 HAVCR2 LAG3 TIGIT TOX TOX2 ENTPD1 LAYN CXCL13 DUSP4 XCL1 XCL2 CD160 CD200 CD226 BTLA GNG4 TNFRSF18 TNFRSF4 MKI67 TOP2A ICOS CXCR6 GZMA GZMB GZMK CCL4 CCL4L2 TNFSF9 TNF IFNG KLRG1 CD28 FOSB EGR1 NR4A1 SELL CCR7 TCF7 LEF1 CX3CR1 PRF1 CMC1 KLRF1 CD244 GNLY FCGR3A TYROBP CD3E CD4 CD8A TRAC TRDC KLRB1
Features
Percent Expressed
0
25
50
75
Average Expression
2
1
0
−1
−2

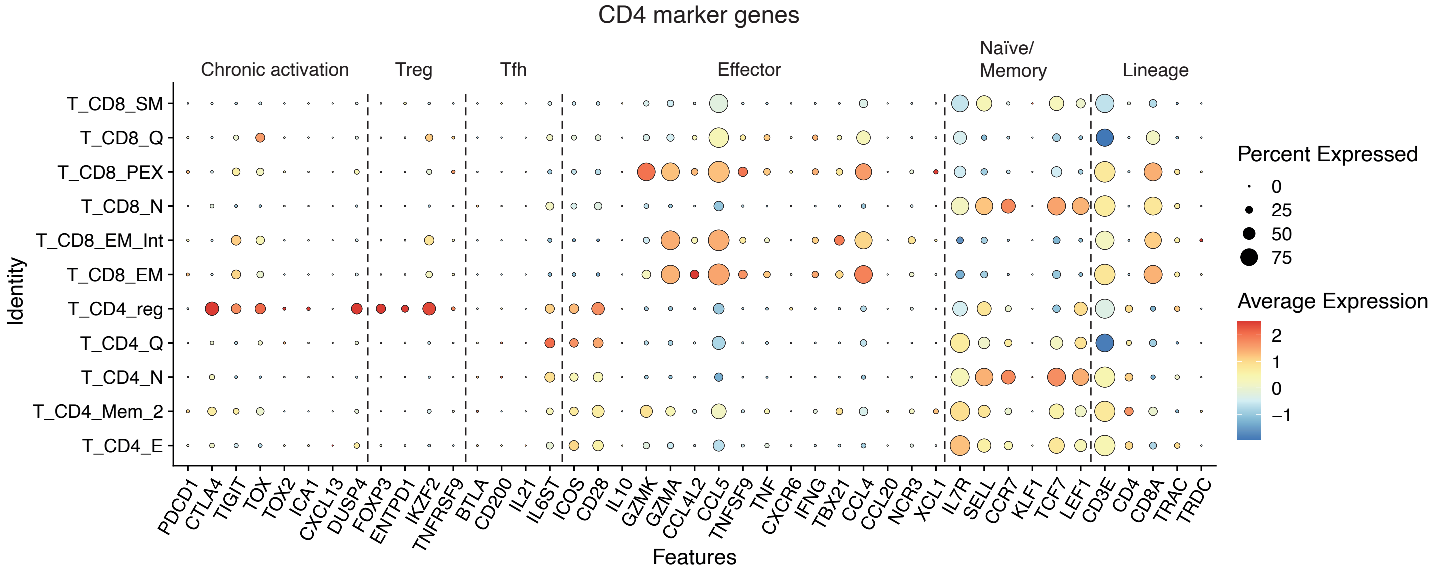
CD4 marker genes
Chronic activation
Treg
Tfh
Effector
Naïve/
Memory
Lineage
T_CD8_SM
T_CD8_Q
T_CD8_PEX
T_CD8_N
T_CD8_EM_Int
T_CD8_EM
T_CD4_reg
T_CD4_Q
T_CD4_N
T_CD4_Mem_2
T_CD4_E
Identity
PDCD1 CTLA4 TIGIT TOX TOX2 ICA1 CXCL13 DUSP4 FOXP3 ENTPD1 IKZF2 TNFRSF9 BTLA CD200 IL21 IL6ST ICOS CD28 IL10 GZMK GZMA CCL4L2 CCL5 TNFSF9 TNF CXCR6 IFNG TBX21 CCL4 CCL20 NCR3 XCL1 IL7R SELL CCR7 KLF1 TCF7 LEF1 CD3E CD4 CD8A TRAC TRDC
Features
Percent Expressed
0
25
50
75
Average Expression
2
1
0
−1

**Supplementary Figure S7. Single-cell transcriptomic landscape of bone marrow T cells in Talquetamab-DRd–treated patients.**

**(A)** UMAP embedding of bone marrow T cells from patients treated with Talquetamab-DRd. UMAP visualization displaying transcriptomic clustering of bone marrow T cells, with overlaid RNA expression used to annotate canonical T cell populations.

**(B–C)** Canonical marker gene expression across T cell subsets. Dot plots showing average expression and detection frequency of curated canonical marker genes across $CD8^{+}$ **(B)** and $CD4^{+}$ **(C)** T cell clusters.

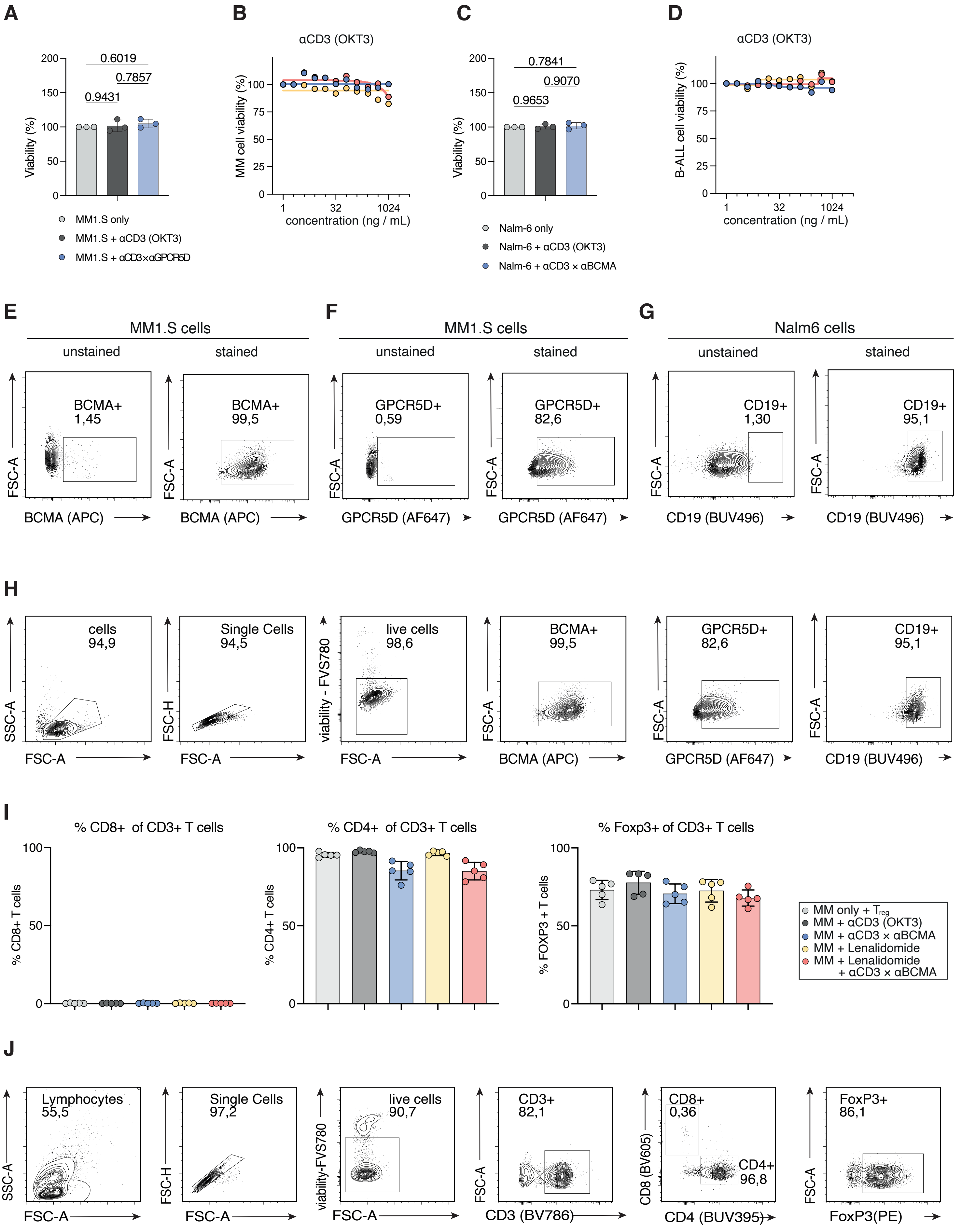

A
Viability (%)
0.6019
0.7857
0.9431
MM1.S only
MM1.S + αCD3 (OKT3)
MM1.S + αCD3×αGPCR5D
B
αCD3 (OKT3)
MM cell viability (%)
concentration (ng / mL)
C
Viability (%)
0.7841
0.9070
0.9653
Nalm-6 only
Nalm-6 + αCD3 (OKT3)
Nalm-6 + αCD3 × αBCMA
D
αCD3 (OKT3)
B-ALL cell viability (%)
concentration (ng / mL)
E
MM1.S cells
unstained
stained
BCMA+ 1,45
BCMA+ 99,5
FSC-A
BCMA (APC)
F
MM1.S cells
unstained
stained
GPCR5D+ 0,59
GPCR5D+ 82,6
GPCR5D (AF647)
G
Nalm6 cells
unstained
stained
CD19+ 1,30
CD19+ 95,1
CD19 (BUV496)
H
cells 94,9
Single Cells 94,5
live cells 98,6
BCMA+ 99,5
GPCR5D+ 82,6
CD19+ 95,1
SSC-A
FSC-H
viability - FVS780
FSC-A
I
% CD8+ of CD3+ T cells
% CD4+ of CD3+ T cells
% Foxp3+ of CD3+ T cells
% CD8+ T cells
% CD4+ T cells
% FOXP3 + T cells
MM only + $T_{reg}$
MM + αCD3 (OKT3)
MM + αCD3 × αBCMA
MM + Lenalidomide
MM + Lenalidomide + αCD3 × αBCMA
J
Lymphocytes 55,5
Single Cells 97,2
live cells 90,7
CD3+ 82,1
CD8+ 0,36
CD4+ 96,8
FoxP3+ 86,1
viability-FVS780
CD3 (BV786)
CD8 (BV605)
CD4 (BUV395)
FoxP3(PE)

**Supplementary Figure S8. Control viability assays and target antigen expression in MM1.S and Nalm6 cells.**

**(A)** Viability of MM1.S cells directly treated for 48-h with equimolar concentrations of Talquetamab (4096 ng/ml) or OKT3 (1585.2 ng/mL) in the absence of T cells. Two-way ANOVA with multiple comparisons (n = 3).

**(B)** Viability of Nalm6 cells directly treated for 48-h with equimolar concentrations of Blinatumomab (1024 ng/ml) or OKT3 (1420.3 ng/mL) in the absence of T cells. Two-way ANOVA with multiple comparisons (n = 3).

**(C)** Viability of MM1.S cells after 48-h coculture with Tregs treated with equimolar concentrations of OKT3 compared to treatment in Figure 4J (n=3).

**(D)** Viability of Nalm6 cells after 48-h coculture with Tregs treated with equimolar concentrations of OKT3 compared to treatment in Figure 4K (n=3).

**(E)** FACS plots showing the expression of BCMA on MM1.S cells.

**(F)** FACS plots showing the expression of GPCR5D on MM1.S cells.

**(G)** FACS plots showing the expression of CD19 on Nalm6 cells.

**(H)** Representative flow cytometric gating used to confirm target marker on cell lines used in coculture assays.

**(I)** Flow cytometric quantification of CD8+, CD4+, and Foxp3+ CD3+ T-cells following 72hr of coculture with MM1S cells and respective treatments.

**(J)** Representative intracellular FACS plots for the gating strategy of CD8+, CD4+ and $FOXP3^+$ expressing cells following 72hr coculture.

**Table 1 | Overview of newly-diagnosed multiple myeloma (NDMM) cohorts – patient and disease characteristics. Related to Figure 1.**

| ID | Cohort | Treatment group | Age[a] | Sex | ECOG | MM subtype | ISS[b] | R-ISS[b] | Cytogenetics | +1q[c] | Soft tissue manifestations |
|---|---|---|---|---|---|---|---|---|---|---|---|
| **Tec-DRd 1** | Majes-TEC-5 | Tec-DRd (Arm A) | 61-65 | Male | 1 | BJ kappa | 3 | 2 | SR | 0 | none |
| **Tec-DRd 2** | Majes-TEC-5 | Tec-DRd (Arm A) | 51-55 | Male | 1 | IgA kappa | 1 | 2 | HR | 0 | none |
| **Tec-DRd 3** | Majes-TEC-5 | Tec-DRd (Arm A) | 56-60 | Male | 0 | IgG lambda | 1 | 1 | SR | 1 | none |
| **Tec-DRd 4** | Majes-TEC-5 | Tec-DRd (Arm A) | 51-55 | Male | 0 | IgG kappa | 2 | 2 | SR | 1 | none |
| **Tec-DRd 5** | Majes-TEC-5 | Tec-DRd (Arm A) | 66-70 | Male | 1 | IgG kappa | 2 | 2 | SR | 0 | none |
| **Tec-DRd 6** | Majes-TEC-5 | Tec-DRd (Arm A1) | 61-65 | Female | 1 | IgG kappa | 1 | 1 | SR | 0 | paramedullary |
| **Tec-DRd 7** | Majes-TEC-5 | Tec-DRd (Arm A1) | 61-65 | Male | 0 | IgA lambda | 2 | 2 | SR | 1 | none |
| **Tec-DRd 8** | Majes-TEC-5 | Tec-DRd (Arm A1) | 61-65 | Female | 0 | IgA lambda | 2 | 2 | SR | 1 | none |
| **Tec-DRd 9** | Majes-TEC-5 | Tec-DRd (Arm A1) | 61-65 | Male | 0 | IgG/A lambda | 1 | 1 | SR | 1 | none |
| **Tec-DRd 10** | Majes-TEC-5 | Tec-DRd (Arm A1) | 61-65 | Male | 0 | IgG kappa | 1 | 1 | SR | 0 | none |
| **Tec-DRd 11** | Majes-TEC-5 | Tec-DRd (Arm A1) | 51-55 | Male | 0 | IgG kappa | 1 | 1 | SR | 0 | paramedullary |
| **Tec-DRD 12** | Majes-TEC-5 | Tec-DRd (Arm D) | 66-70 | Female | 1 | BJ lambda | 2 | 2 | HR | 1 | none |
| **Tec-DRD 13** | Majes-TEC-5 | Tec-DRd (Arm D) | 51-55 | Male | 2 | IgA kappa | 2 | 2 | HR | 1 | none |
| **Tec-DRD 14** | Majes-TEC-5 | Tec-DRd (Arm D) | 56-60 | Male | 0 | BJ kappa | 1 | 2 | HR | 0 | none |
| **Tec-D-VRd 1** | Majes-TEC-5 | Tec-D-VRd (Arm B) | 46-50 | Male | 0 | IgG lambda | 1 | 1 | SR | 1 | none |
| **Tec-D-VRd 2** | Majes-TEC-5 | Tec-D-VRd (Arm B) | 36-40 | Male | 1 | IgG kappa | 1 | 2 | HR | 1 | none |
| **Tec-D-VRd 3** | Majes-TEC-5 | Tec-D-VRd (Arm B) | 56-60 | Male | 1 | IgA kappa | 1 | 1 | SR | 1 | paramedullary |
| **Tal-DRD 1** | Majes-TEC-5 | Tal-DRd (Arm E1) | 46-50 | Female | 0 | IgA lambda | 1 | 2 | HR | 1 | none |
| **Tal-DRD 2** | Majes-TEC-5 | Tal-DRd (Arm E) | 66-70 | Male | 2 | IgG kappa | 1 | 2 | HR | 0 | none |
| **Tal-DRD 3** | Majes-TEC-5 | Tal-DRd (Arm E) | 61-65 | Male | 0 | BJ lambda | 3 | 2 | SR | 1 | none |
| **D-Rd 1** | Standard-of-care | DRd | 81-85 | Male | 0 | BJ kappa | 3 | 2 | SR | 0 | paramedullary |
| **D-Rd 2** | Standard-of-care | DRd | 71-75 | Male | 2 | IgG kappa | 1 | 1 | SR | 0 | none |
| **D-Rd 3** | Standard-of-care | DRd | 71-75 | Male | NA | BJ kappa | 1 | 1 | SR | 0 | paramedullary |
| **D-Rd 4** | Standard-of-care | DRd | 81-85 | Male | NA | IgG lambda | 2 | 2 | SR | 0 | none |
| **D-Rd 5** | Standard-of-care | DRd | 66-70 | Female | NA | IgG kappa | 2 | 2 | SR | 0 | none |
| **D-Rd 6** | Standard-of-care | DRd | 71-75 | Female | 1 | IgG kappa | 2 | 2 | HR | 1 | extramedullary |

a. Determined at cycle 1 day 1. b. Last available assessment prior to treatment initiation. c. Chromosome 1q21 abnormalities including 1q gain (3 copies) or 1q amplification (≥ 4 copies). BJ, Bence Jones. DRd, Daratumumab/Lenalidomide/Dexamethasone. ECOG, Eastern Cooperative Oncology Group performance status. HR, high risk (del(17p), t(4;14) and/or t(14;16). ISS, International Staging System. MM, multiple myeloma. NA, not available. R-ISS, Revised International Staging System. SR, standard risk. Tal-DRd, Talquetamab/Daratumumab/Lenalidomide/Dexamethasone. Tec-D-VRd, Teclistamab/Daratumumab/Bortezomib/Lenalidomide/Dexamethasone. Tec-DRd, Teclistamab/Daratumumab/Lenalidomide/Dexamethasone.
